# Generalizable CT Vision-Language Modeling for Population Health and Disease Risk

**DOI:** 10.1101/2025.07.03.25330654

**Authors:** Cameron A. Beeche, Joonghyun Kim, Hamed Tavolinejad, Bingxin Zhao, Jessie Dong, Rakesh Sharma, Jeffrey Duda, James Gee, Farouk Dako, Anurag Verma, Colleen Morse, Bojian Hou, Li Shen, Hersh Sagreiya, Christos Davatzikos, Scott Damrauer, Rohan Shad, Marylyn D. Ritchie, Daniel Rader, Qi Long, Eric Eaton, Gengwei Zhang, Tianlong Chen, Charles E. Kahn, Julio Chirinos, Walter R. Witschey, Penn Medicine Biobank

## Abstract

Vision-language foundation models (VLMs) for computed tomography (CT) are emerging tools that learn generalizable representations from large-scale clinical imaging data. While these models can predict task-specific labels, the extent to which their representations capture the clinical, physiological, and longitudinal variation of real-world patient populations remains unclear. We introduce Percival, a CT-native VLM trained on more than 400,000 CT-report pairs from the Penn Medicine BioBank using a dual-encoder symmetric contrastive framework. Across over 20,000 held-out participants, Percival’s latent space aligns with demographic, physiological, and laboratory variation, and supports phenome-wide associations across the electronic health record. We evaluate Percival against alternative foundation-model paradigms, including vision-only contrastive and multi-organ segmentation, demonstrating that vision-language pretraining captures clinical information not fully accessible to vision-only alternatives across disease classification and longitudinal risk modeling. Together, these findings indicate that CT-VLMs uncover latent structure aligned with clinical, physiological, and prognostic variation across the disease-prevalence spectrum.

## INTRODUCTION

Computed tomography (CT) is widely used for disease diagnosis and monitoring^1–3^, but rising utilization, radiology workforce shortages, and access disparities^4–6^ are driving the need for AI-powered tools to improve efficiency and support precision medicine^7–14^. Self-supervised vision-language models (VLMs), trained using CT volumes and accompanying radiology reports, have been proposed to address these issues when fine-tuned for downstream tasks such as disease classification, and report drafting^15,16^. However, it remains unclear how well these models capture the broader clinical and physiological variation present in real-world patient populations, including conditions whose primary diagnosis does not rely on CT. Existing evaluations offer limited insight into whether CT-VLMs learn representations that align with patient physiology, laboratory biomarkers, multisystem disease burden, or longitudinal risk. Moreover, CT-VLMs have not been systematically characterized at biobank scale or benchmarked against alternative foundation-model paradigms that operate on the three-dimensional computed tomography. Clarifying these properties is important for understanding what CT-VLM representations capture about patient populations and for assessing their value as foundation models for medical imaging.

Leveraging the Penn Medicine BioBank (PMBB), we developed Percival, a vision-language foundation model for three-dimensional CT imaging. Percival was trained on more than 400,000 CT volumes paired with corresponding radiology reports collected from multiple hospitals spanning the Penn Medicine Healthcare system, capturing substantial diversity in imaging protocols, scanner types, contrast administration, and patient populations. The PMBB additionally provides linked laboratory measurements, structured physiological data, and longitudinal electronic health records, enabling biobank-scale characterization of how a pretrained CT-VLM’s latent representations align with measured clinical, physiological, and prognostic variables. This integration of imaging with longitudinal patient data extends beyond what typical CT-VLM training cohorts support, which generally provide imaging and reports but limited linked clinical follow-up.

In this study, we sought to characterize how a CT-based VLM’s latent representations align with measured clinical, physiological, and longitudinal variables within a large clinical biobank. First, we quantified associations between Percival’s latent representations and demographic factors, body-size measures, and laboratory biomarkers, and tested whether these embeddings support synthetic estimation of laboratory values directly from imaging. Second, we mapped phenome-wide associations between Percival’s latent representations and clinical diagnoses, characterized correspondence with multisystem disease burden, and benchmarked classification performance against alternative foundation-model paradigms across the disease-prevalence spectrum. Finally, we compared how the feature representations of these foundation-model paradigms encode longitudinal disease risk using Cox proportional-hazards modeling. An overview of the study design is presented in **Figure 1**. To support continued research, we publicly release Percival’s pretrained weights and accompanying code (https://github.com/cams2b/percival).

**Figure 1.**
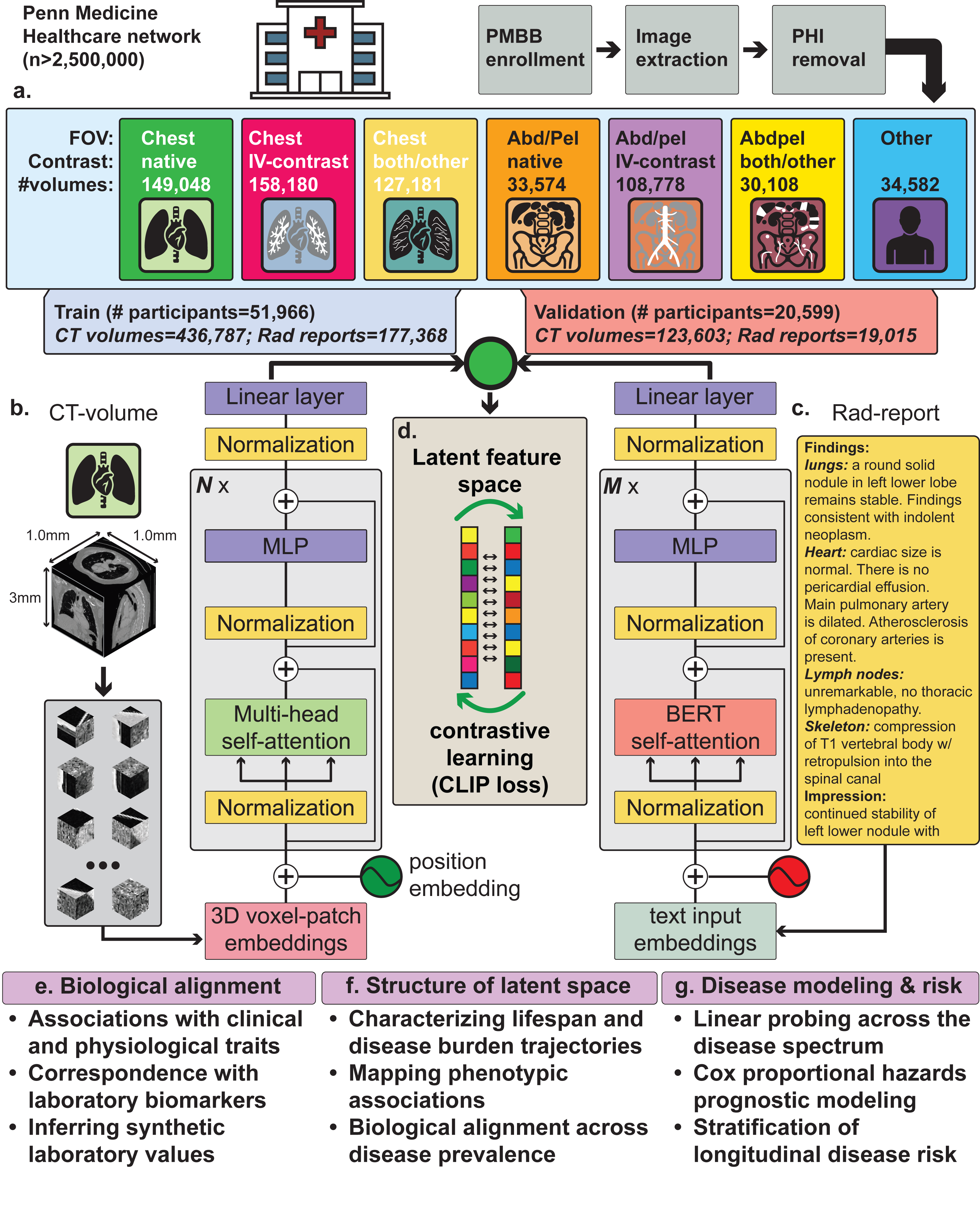
Overview of data, model design, and downstream analyses. (**a**) Overview of the Penn Medicine Biobank (PMBB) computed tomography (CT) imaging data used in this study, stratified by anatomical region and intravenous contrast administration, and the participant-level train-validation split. (**b**) Percival’s vision encoder, an augmented vision transformer (ViT) supporting three-dimensional patch embeddings with a 16 x 16 x 8 voxel patch size. (**c**) Percival’s language encoder, a pretrained CXR-BERT specialized architecture. (**d**) Volumetric CT imaging data and radiology report text are aligned in a 768-dimensional latent feature space using a symmetric contrastive (InfoNCE) loss function. (**e**) Evaluation of Percival’s biological alignment through associations with clinical and physiological traits, laboratory biomarkers, and synthetic laboratory value prediction. (**f**) Characterization of Percival’s latent feature space through lifespan disease burden trajectories, phenotypic associations, and disease-prevalence analyses. (**g**) Disease modeling and risk through linear probing for classification and Cox proportional-hazards prognostic modeling, with longitudinal risk stratification.

## RESULTS

Here, we present Percival, a contrastive vision-language model for three-dimensional CT imaging trained in a manner consistent with recent CT-VLM efforts such as Merlin and CT-CLIP, pairing volumetric CT studies with their accompanying radiology reports to learn shared visual-textual representations^15,16^. The study cohort included 72,565 PMBB participants (51,966 for training and 20,599 for validation) with comparable demographic distributions (**Table S1**). The training subset contributed 436,787 CT volumes spanning diverse anatomical regions, contrast phases, scanner types, and acquisition protocols across multiple clinical sites (**Tables S2**).

Throughout this manuscript, “Percival” refers to the 86M-parameter ViT-Base configuration; the Percival-Tiny (6M) and Percival-Small (22M) variants are released alongside the manuscript. To verify Percival’s out-of-domain generalization, we evaluated zero-shot and linear probing classification on the external CT-RATE dataset, where the model achieved performance comparable to in-domain CT-CLIP^17^ (**Figs. S1-S2, Table S3**). Unsupervised visualization of the learned representations further revealed organized clustering by imaging acquisition attributes (**Fig. S3**), including field of view, intravenous contrast administration, and anatomical region, as well as by participant demographic attributes including age, sex, and BMI (**Fig. S4**), indicating that Percival captures meaningful variation present in real-world imaging data.

### Biological associations with Percival’s latent space

Principal component analysis applied to Percival’s latent feature embeddings yielded a gradual decay of variance across the leading components (**Fig. S5**). Linear regression analyses testing associations between these components and nine continuous physiological traits (**Methods**), identified 67 of 90 PC-trait associations as significant after Bonferroni correction (*P*<0.05/90; **Table S4**). Age was most strongly associated with PC2 (R^2^=0.35; **Fig. 2A**), whereas BMI was most strongly associated with PC9 (R^2^=0.11; **Fig. 2C**). Height was most strongly associated with PC7 (R^2^=0.14). Blood pressure, pulse, oxygen saturation, and respiratory rate showed statistically significant but smaller associations across multiple PCs (R^2^<0.025), indicating limited representation of these vital-sign traits in the leading PCs of the latent space. Notably, demographic and anthropometric signal was distributed across multiple components rather than concentrated on a single dominant axis, with each trait showing meaningful contributions from three or more PCs. Sex differences were evaluated using ANOVA with effect size measured by Cohen’s D (**Methods**; **Fig. 2B**; **Table S5**). All ten PCs showed Bonferroni-significant sex effects. The leading components (PC1-PC4) demonstrated moderate effect sizes (|Cohen’s D| < 0.5), suggesting that these latent features encode shared physiological variation rather than sex-specific patterns. In contrast, PC7 demonstrated a large effect size (|Cohen’s D| = 1.08), consistent with the latent space capturing morphological variation between males and females (**Fig. S4b**).

**Figure 2.**
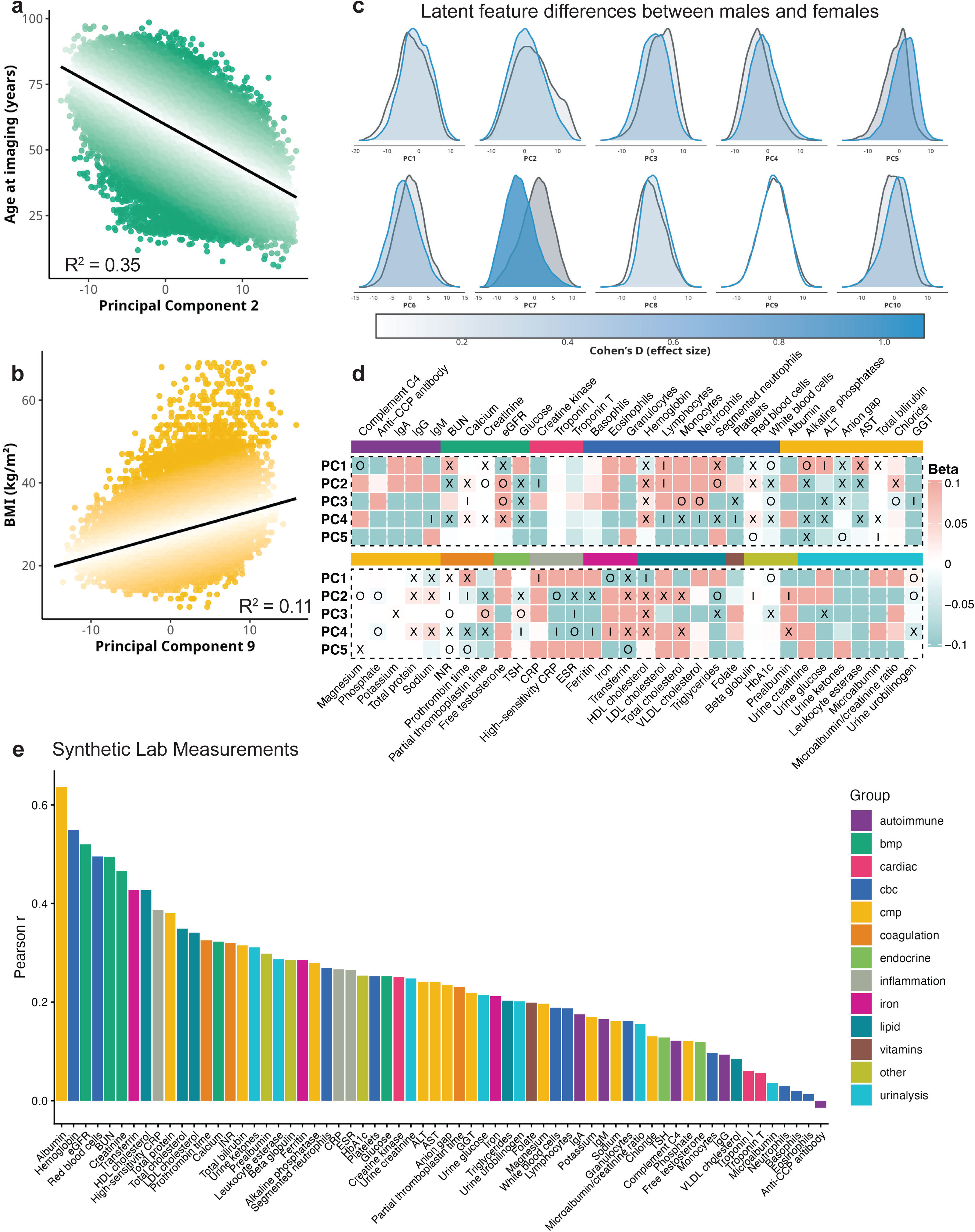
Biological associations of Percival’s latent space. (**a**) Association between participant age at imaging and latent principal component 2 (PC2) shown using linear regression (R^2^=0.35). (**b**) Density distribution of latent PCs 1-10 stratified by participant sex. Color intensity reflects the magnitude of Cohen’s D effect size for male-female separation. (**c**) Association between body mass index (BMI) and latent principal component 9 (PC9) shown using linear regression (R^2^=0.11). (**d**) Half-half discovery–replication laboratory-wide association study (LabWAS) for latent PCs 1-5, organized by laboratory group. An “X” indicates associations reaching Bonferroni significance in both discovery and replication cohorts with directionally concordant effects; “I” and “O” indicate Bonferroni significance in the discovery and replication cohorts only, respectively. (**e**) Pearson correlation coefficients between predicted and observed laboratory values from synthetic laboratory prediction models trained on Percival’s latent features, ranked by performance and colored by laboratory group.

We then conducted a laboratory-wide association study (LabWAS) using a half-half discovery-replication design to uncover the biological associations between principal components (PCs 1-10) derived from Percival’s latent feature embeddings (**Fig. 2D**; **Table S6**). Following Bonferroni correction for multiple comparisons (*P*<0.05/640), our discovery cohort (average n=2,652) identified 153 significant associations with 50 laboratory measurements, with 113 associations across 39 lab measurements being further verified in the replication cohort (average n=2,606). Moreover, all associations that passed significance among both the discovery-replication cohorts were concordant and in the same direction (**Fig. S6**). Percival’s latent feature space demonstrated consistent associations with laboratory markers of renal function, including blood urea nitrogen (BUN), creatinine, and estimated glomerular filtration rate (eGFR). These markers reflect underlying glomerular and tubular function and are frequently used to assess kidney health^18–20^. Several components showed significant associations with hepatic and biliary-related lab measurements, including alkaline phosphatase (ALP), alanine aminotransferase (ALT), aspartate aminotransferase (AST), total bilirubin, and urobilinogen, which serve as primary indicators of hepatocellular injury, cholestasis, and bile metabolism^21,22^. Replicated associations were also observed with metabolic and lipid markers, including glucose, HbA1c, total cholesterol, HDL, LDL, and triglycerides, reflecting glycemic control and lipid regulation^23–27^. Hematologic associations included hemoglobin, red and white blood cell counts, platelets, and differential leukocyte fractions, suggesting that the latent space captures variation related to hematopoietic status. Additional replicated associations were observed for inflammatory markers (high-sensitivity CRP, ESR), coagulation markers (PT, PTT, INR), iron-related markers (ferritin, transferrin), and electrolyte and acid-base markers (sodium, potassium, chloride, calcium, magnesium, anion gap). CRP and ESR are established biomarkers of systemic inflammation and have been consistently linked to cardiovascular disease risk and atherosclerosis^28,29^.

### Synthetic lab values

We next generated synthetic laboratory values from Percival’s latent feature space to assess the extent to which CT-VLM’s feature representations capture biochemical variation. Linear probing of Percival’s latent feature space was performed using a 3-layer multilayer perceptron with 5-fold subject-grouped cross-validation, allowing estimation of laboratory traits in participants without concurrent blood draws (**Fig. 2E**). This approach provides an in-silico approximation of physiological markers directly from CT-derived embeddings. Across 64 laboratory measurements, overall predictive performance was weak (mean cross-validated R^2^=0.04), with the embedding showing variable correspondence to different biomarker classes. Several laboratory traits showed substantially stronger correspondence with the latent space, including albumin (R^2^=0.40), hemoglobin (R^2^=0.30), estimated glomerular filtration rate (R^2^=0.26), red blood cell count (R^2^=0.24), blood urea nitrogen (R^2^=0.24), and creatinine (R^2^=0.21). At a group level, the basic metabolic panel showed the strongest aggregate performance (mean R^2^=0.17), while autoimmune, endocrine, and urinalysis markers were not meaningfully recovered from the embedding. **Figure 2E** presents representative performance across laboratory panels, with complete results summarized in **Table S7**.

### Disease enrichment patterns and lifespan disease burden

To characterize the breadth of disease-related variation represented in Percival’s latent space, we conducted a half-half discovery (average n=10,297) replication (average n=10,296) phenome-wide association study (PheWAS) across 588 phecodes in the PMBB validation cohort. This analysis revealed extensive phenotypic alignment between Percival’s embeddings and clinical diagnoses, with 1,022 associations across 349 phecodes reaching Bonferroni significance (*P*<8.50x10^-6^) in both cohorts with directionally concordant effects (**Fig. 3A, Table S8**). These associations spanned multiple organ systems—including 68 circulatory, 41 respiratory, 24 neoplastic, and 42 endocrine/metabolic conditions—suggesting that Percival’s latent representations capture phenotypic variation extending beyond the findings typically described on CT. Sensitivity analyses restricting the imaging-to-diagnosis window to 30, 15, and 7 days yielded 230 phecodes reaching phenome-wide significance across all four windows (**Fig. S7, Table S9**).

**Figure 3.**
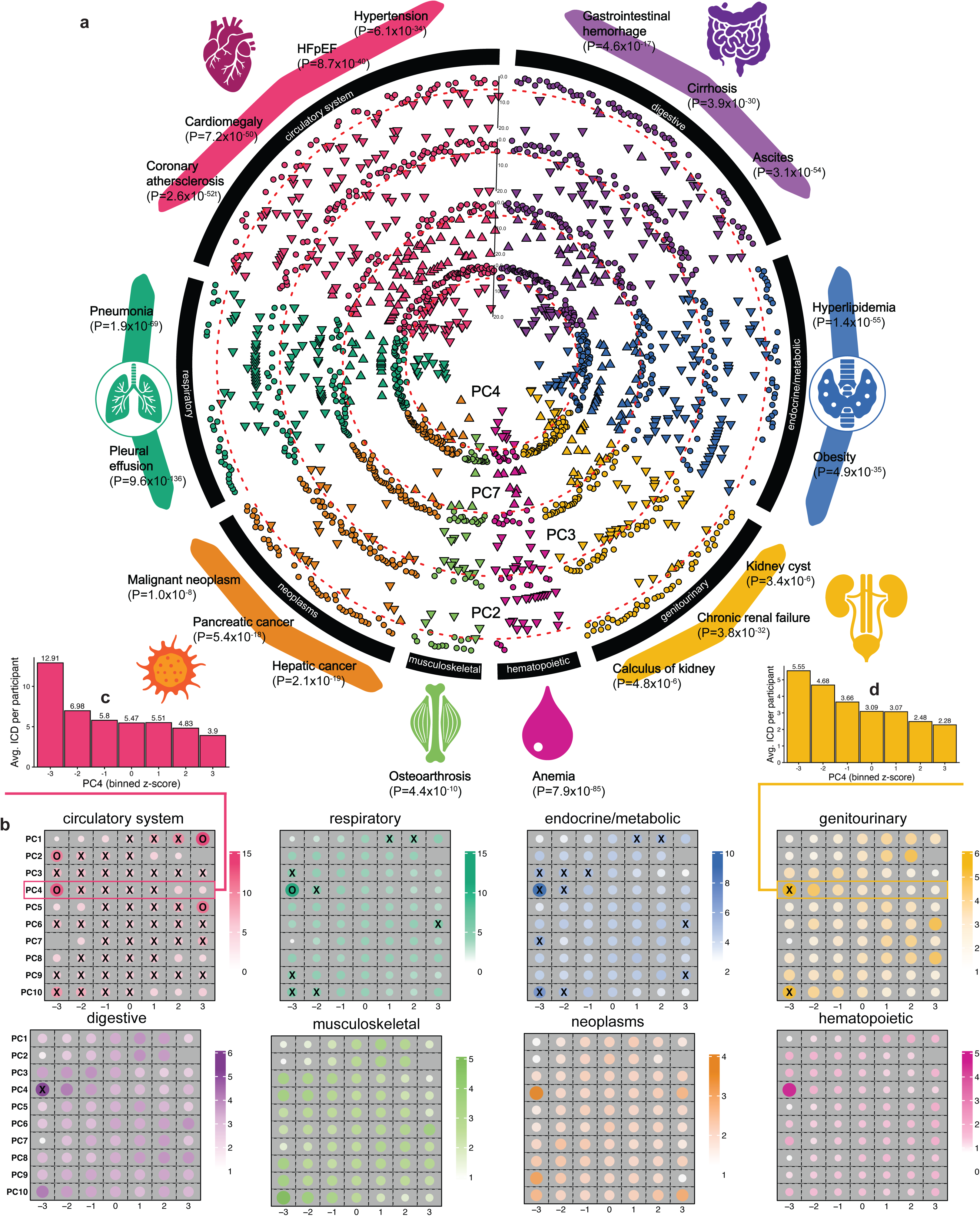
Phenome-wide disease enrichment and lifespan disease burden associations with Percival’s latent feature space. (**a**) Circular Manhattan plot depicting phenome-wide associations between the top four most associated principal components of Percival’s latent feature space and clinical diagnoses across eight disease groups (circulatory system, respiratory, digestive, genitourinary, musculoskeletal, neoplasms, endocrine/metabolic, hematopoietic) in the discovery cohort of a 50-50 discovery-replication design. The red dashed line marks the discovery-cohort Bonferroni significance threshold. Upward triangles indicate positive associations with disease prevalence; downward triangles indicate negative associations. (**b**) Lifespan disease-burden mapping for z-score-normalized latent PCs 1-10, computed in the PMBB validation cohort using one CT volume per participant (earliest scan by age at imaging). Each of the eight disease-group panels shows the mean number of post-imaging conditions accumulated per participant within each (PC, z-score bin) cell, with z-scores binned to integers in the range [-3, +3]. An “X” indicates cells with a mean of 5 to 10 conditions per participant; an “O” indicates cells with a mean exceeding 10 conditions per participant. (**c**) Bar plot illustration of circulatory disease burden for component 4. (**d**) Bar plot illustration of genitourinary disease burden for component 4.

To assess the longitudinal clinical impact of these phenome-wide associations, we next quantified post-imaging disease accumulation as a function of latent biological features. Each participant’s principal component axes were binned using standardized z-score (−3 to +3). For each bin, we measured the mean number of unique conditions recorded after the imaging date, defining a prospective measure of lifespan disease burden (**Fig. 3B, Table S10**). A clear disease accumulation gradient was observed across multiple components, with individuals at the extremes of PC values demonstrating a markedly greater accumulation of new diagnoses across circulatory, metabolic/endocrine, and respiratory systems. Specifically, component 4 exhibited the strongest disease gradient for circulatory diseases, with participants binned at <-3 accruing on average 12.9 circulatory related diseases post imaging (**Fig. 3C**). Similar patterns were observed amongst component 4 for endocrine and metabolic diseases, with participants binned at <-3 accruing >5 diseases over their lifespan (**Fig. 3D**). These relationships suggest that the latent space of a CT-VLM can capture multisystem patterns of disease risk and phenotypic variation.

### Disease classification

To characterize Percival’s latent capability for disease classification and compare it to alternative, non-VLM, foundation-model designs, we benchmarked performance against two demographic baselines (age + sex; age + sex + number of prior imaging studies), CT-FM^30^ (a vision-only contrastive foundation model), and TotalSegmentator^13^ (a deterministic organ-level segmentation model). Specifically, we applied linear probing across clinical disease states in a region-specific manner, separately for chest (702 phecodes) and abdomen/pelvis (624 phecodes) CT volumes. The DeLong test was used to assess whether Percival’s discriminative performance exceeded that of comparator models^31^. The number of positive cases per condition varied widely, with 50% including fewer than 500 positive cases (**Fig. 4a, Fig. S8a**). When performance was examined as a function of disease prevalence, AUC exhibited a modest, positive relationship (**Fig. 4b, Fig. S8b**). Across the chest and abdomen/pelvis classification analyses, Percival’s latent feature space provided discriminative signal for a broad range of clinical conditions (**Fig. 4c**). In chest CT, Percival significantly outperformed the demographic baseline on 555 conditions, the demographic baseline adjusted for prior imaging studies on 544 conditions, CT-FM on 512, and TotalSegmentator on 315 following FDR correction at the 5% threshold. In abdomen/pelvis CT, Percival significantly outperformed the demographic baseline on 511 conditions, the demographic baseline adjusted for prior imaging studies on 500 conditions, CT-FM on 442, and TotalSegmentator on 272 conditions. The reverse comparisons were markedly less frequent. In chest CT, the demographic baseline outperformed Percival on 10 conditions, the demographic baseline adjusted for prior imaging studies on 6, TotalSegmentator on 9, and CT-FM on 1. In abdomen/pelvis CT, the corresponding counts were 6, 4, 5, and 0. In total, Percival outperformed all comparator models on 279 conditions in chest CT and 230 conditions in abdomen/pelvis CT (**Table S11-S12**). Percival additionally demonstrated the strongest calibration profile across both anatomical regions, exceeding all comparator models (**Figs. S9-S10**, **Table S11-S12**). Specifically, Percival demonstrated significant performance gains across all comparator models for 59 circulatory disease states in chest CT (mean AUC=0.84; range=0.65, 0.98) and 33 in abdomen/pelvis CT (mean AUC=0.84; range=0.65, 0.94). Respiratory diseases demonstrated similar trends, with Percival showing significant performance gains across all comparator models for 43 conditions in chest CT (mean AUC=0.83; range=0.67, 0.97) and 19 in abdomen/pelvis CT (mean AUC=0.84; range=0.71, 0.96). Neoplastic diseases showed a similar pattern, with Percival demonstrating significant performance gains across all comparator models for 38 conditions in chest CT (mean AUC=0.84; range=0.71, 0.94) and 25 in abdomen/pelvis CT (mean AUC=0.81; range=0.62, 0.91).

**Figure 4.**
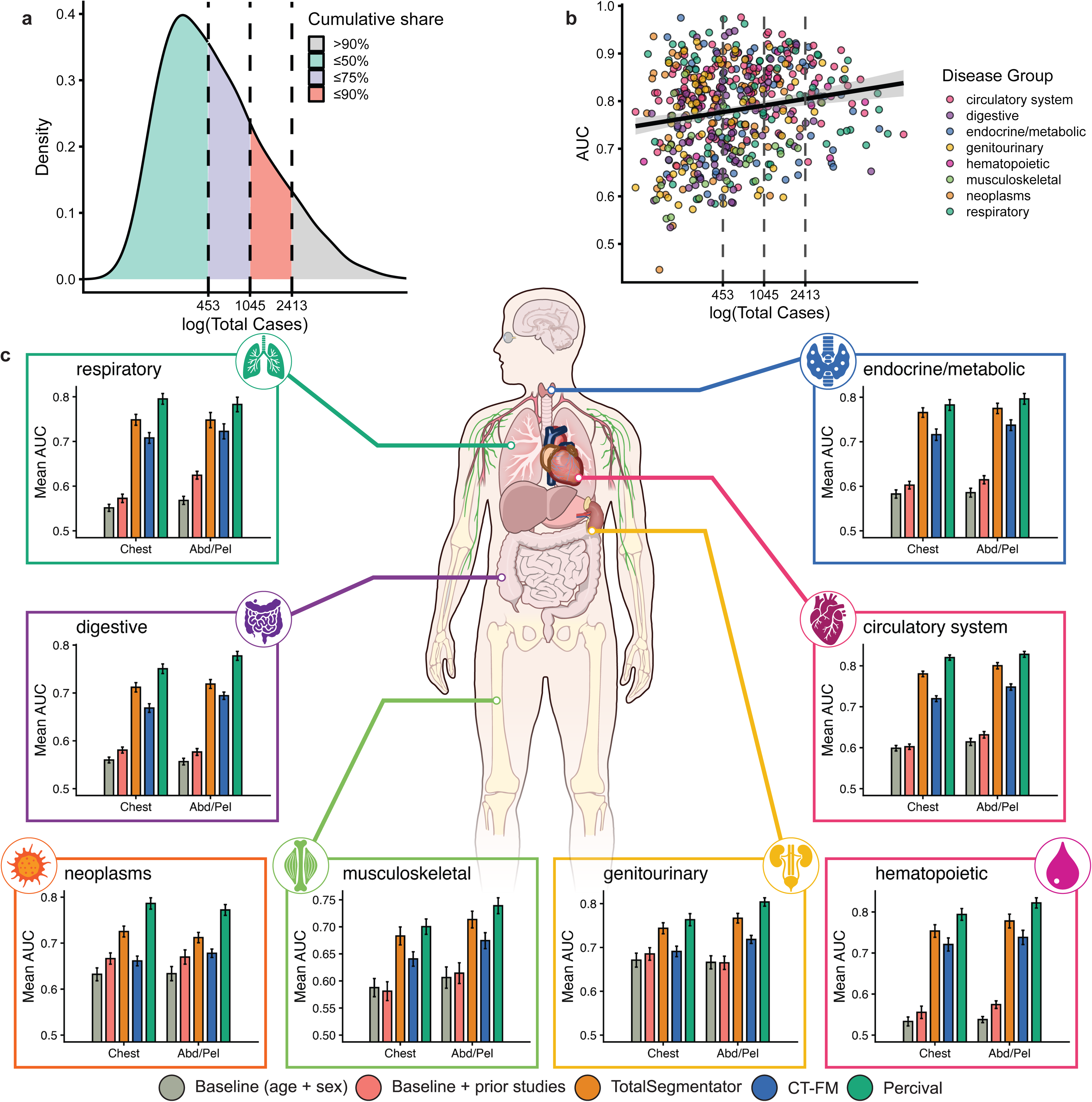
Disease classification performance across Percival’s latent feature space. Using ICD-10CM codes within 60 days of imaging, were used to evaluate Percival’s ability to perform disease classification, stratified by chest and abdomen/pelvis CT volumes and benchmarked against the demographic baseline (age + sex), the baseline + prior studies (referring to the number of previous studies accumulated up to the point of imaging), CT-FM, and TotalSegmentator. **(a)** Density distribution of the number of positive disease cases (log-transformed) across all chest CT classification tasks, shaded by cumulative share quartiles (≤50%, ≤75%, ≤90%, >90%). **(b)** Validation-set area under the receiver operating characteristic curve (AUC) for each chest CT classification task plotted against the log-transformed number of positive cases, colored by disease group. **(c)** Mean validation-set AUC across the eight disease groups (circulatory system, respiratory, digestive, genitourinary, musculoskeletal, neoplasms, endocrine/metabolic, hematopoietic) for chest and abdomen/pelvis CT, comparing Percival to the demographic baseline (age + sex), TotalSegmentator, and CT-FM. Error bars represent the standard error of the mean across conditions within each disease group. Human anatomical art provided by NIH BIOART^48^.

Additional improvements were identified in chest CT across digestive (33, mean AUC=0.81), genitourinary (8, mean AUC=0.87), endocrine/metabolic (25, mean AUC=0.83), and musculoskeletal (3, mean AUC=0.79) conditions, with corresponding gains in abdomen/pelvis CT for digestive (52, mean AUC=0.82), genitourinary (20, mean AUC=0.82), endocrine/metabolic (25, mean AUC=0.85), and musculoskeletal (4, mean AUC=0.77) conditions. Sex-specific replication of disease classification was assessed across 404 chest and 364 abdomen/pelvis conditions. Percival’s cross-sex concordance matched TotalSegmentator on chest CT (R=0.85) and was highest on abdomen/pelvis CT (R=0.83), exceeding TotalSegmentator (R=0.80), CT-FM (R=0.78), and the demographic baseline (R=0.30) (**Fig. S11-S12; Table S13-S14**).

### Examination of outcomes across the disease spectrum

We next examined a curated set of disease states spanning multiple organ systems and varying levels of CT indication to characterize Percival’s discriminative performance across conditions with established CT correlates and those whose primary diagnosis does not rely on CT (**Fig. 5**). The performance margin between Percival and TotalSegmentator was narrowest for disease states characterized by intensity and attenuation changes, such as vascular calcification in atherosclerosis and mass-forming malignancy, resulting in attenuation differences. Specifically, when examining coronary atherosclerosis on chest CT, Percival achieved an AUC of 0.85 (95% CI=0.84, 0.85) compared to TotalSegmentator’s 0.79 (95% CI=0.78, 0.80) (**Fig. 5c**). For pancreatic cancer, Percival achieved an AUC of 0.91 (95% CI=0.90, 0.93) vs TotalSegmentator’s 0.84 (95% CI=0.82, 0.86) (**Fig. 5i**). Similar patterns were observed for bronchiectasis, bacterial pneumonia, and cirrhosis, where volume and attenuation changes similarly narrowed the gap between models (**Figs. 5d, 5e, 5l**). For standard pneumonia, TotalSegmentator and CT-FM exhibited comparable performance, whereas Percival’s performance margin widened (**Fig. 5f**). Percival also showed substantial performance gains over both comparator models for lung cancer (**Fig. 5g**) and acute lymphoid leukemia (**Fig. 5h**), with the gap being widest for lung cancer. For disease states with subtle imaging manifestations or whose primary diagnosis does not rely on CT, Percival maintained strong predictive performance. Atrial fibrillation reached an AUC of 0.84 (95% CI=0.83, 0.85) on chest CT (**Fig. 5a**), whereas heart failure with preserved ejection fraction (HFpEF) reached an AUC of 0.91 (95% CI=0.89, 0.92) (**Fig. 5b**). Strong predictive signal was further observed for conditions not typically assessed on CT, including chronic renal failure and iron deficiency anemia (**Figs. 5j-5k**).

**Figure 5.**
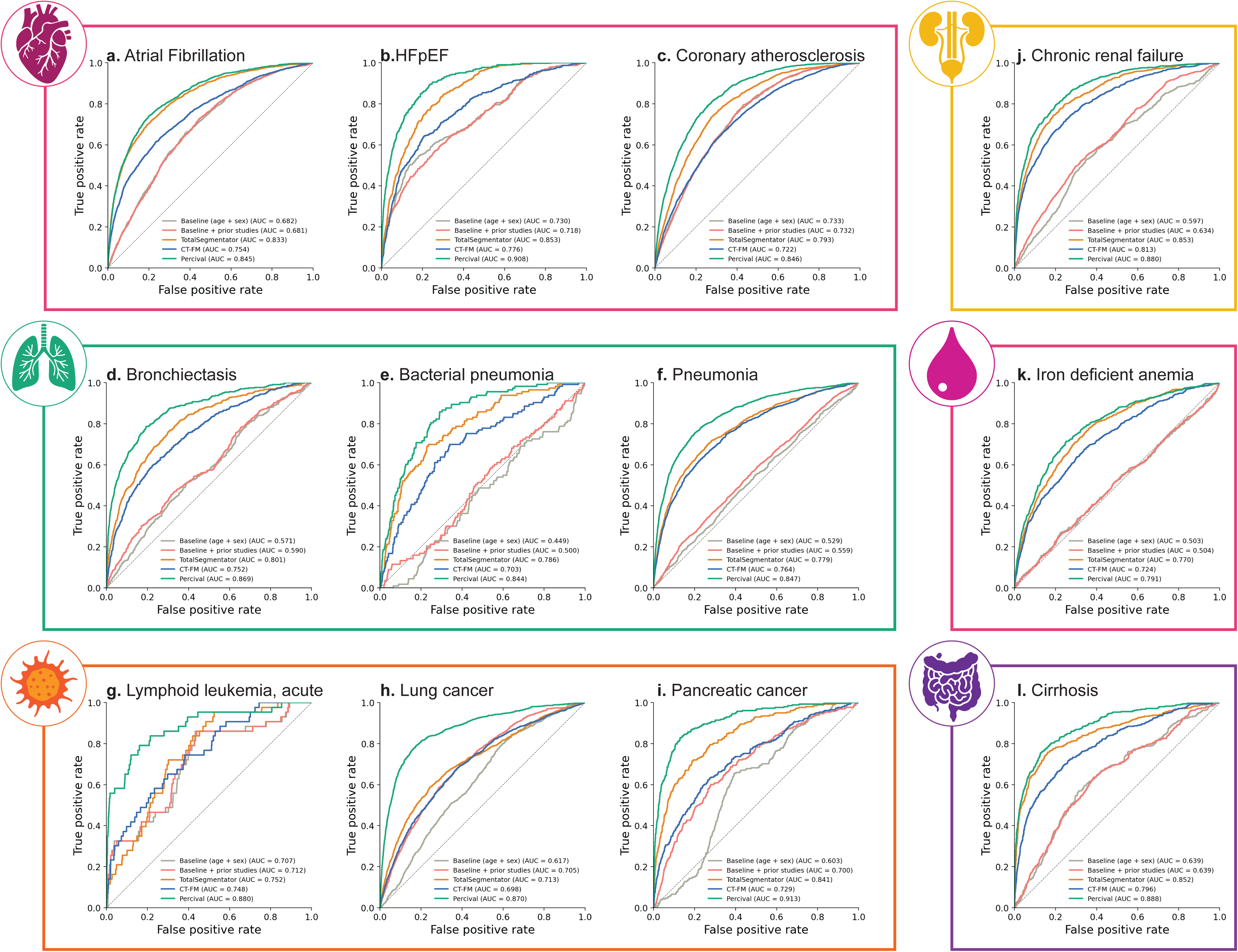
Disease classification performance across the disease spectrum. Receiver operating characteristic curves for Percival, TotalSegmentator, CT-FM, and the demographic baseline (age + sex), and the baseline + prior studies (referring to all previous studies accumulated up to the point of imaging), across twelve representative clinical conditions spanning six disease groups. Each panel reports the validation-set area under the receiver operating characteristic curve (AUC) for the four models. (**a**) Atrial fibrillation. (**b**) Heart failure with preserved ejection fraction (HFpEF). (**c**) Coronary atherosclerosis. (**d**) Bronchiectasis. (**e**) Bacterial pneumonia. (**f**) Pneumonia. (**g**) Lymphoid leukemia, acute. (**h**) Lung cancer. (**i**) Pancreatic cancer. (**j**) Chronic renal failure. (**k**) Iron deficiency anemia. (**l**) Cirrhosis.

### Prognostic modeling

To evaluate whether Percival’s pretrained representations capture information relevant to future health trajectories, we examined Percival’s prognostic utility across a wide range of clinical outcomes using ridge-regularized Cox proportional-hazards models, stratified by chest and abdomen/pelvis CT volumes (**Fig. 6**). Our analysis proceeded in two parts. First, we applied Cox proportional-hazards models across 1,059 conditions in chest CT and 957 conditions in abdomen/pelvis CT, quantifying prognostic signal using the concordance index (C-index). Second, for each condition, we derived latent feature-based risk stratification profiles that separated participants into low-risk (bottom 80%) and high-risk (top 20%) groups, with differences in event-free survival evaluated using the log-rank test with significance determined by Bonferroni correction (**Figs. 6c-h**).

**Figure 6.**
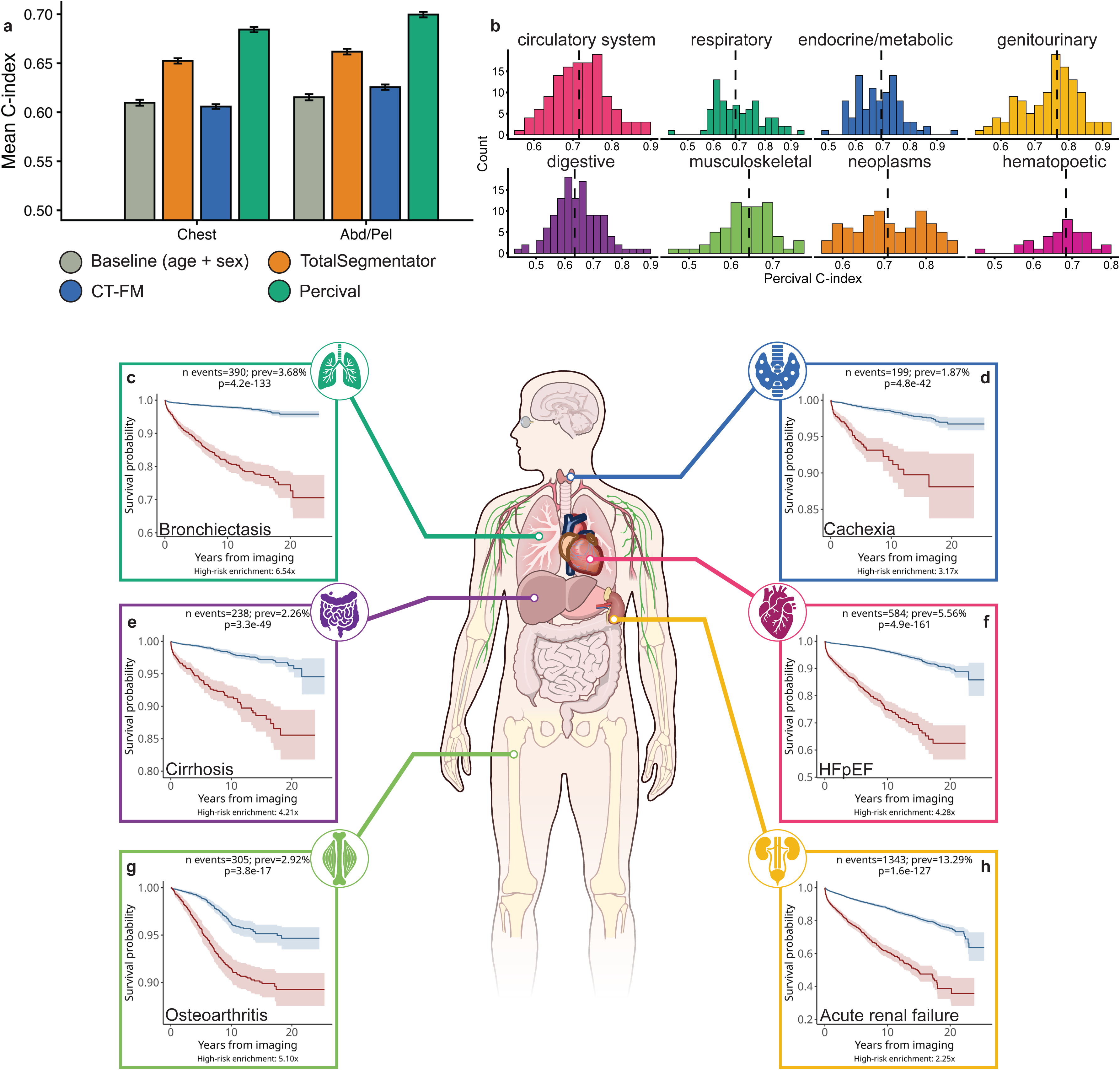
Prognostic utility of Percival’s latent feature space using ridge-regularized Cox proportional-hazards modeling. (**a**) Mean concordance index (C-index) across all conditions for Percival, TotalSegmentator, CT-FM, and the demographic baseline (age + sex), shown separately for chest and abdomen/pelvis CT. Error bars represent the standard error of the mean across conditions. (**b**) Distribution of Percival’s C-index across conditions within each of the eight disease groups (circulatory system, respiratory, endocrine/metabolic, genitourinary, digestive, musculoskeletal, neoplasms, hematopoietic) for chest CT. Dashed vertical lines mark the median C-index per disease group. (**c-h**) Kaplan-Meier survival curves illustrating Percival’s risk stratification for six representative clinical outcomes on chest CT: (**c**) bronchiectasis (respiratory), (**d**) cachexia (endocrine/metabolic), (**e**) cirrhosis (digestive), (**f**) heart failure with preserved ejection fraction (HFpEF; circulatory), (**g**) osteoarthritis (musculoskeletal), and (**h**) acute renal failure (genitourinary). Participants were stratified into low-risk (bottom 80%, blue) and high-risk (top 20%, red) groups based on the linear predictor from the fitted Cox model. Each KM panel reports the number of events, prevalence, log-rank P-value, and high-risk hazard enrichment. Human anatomical art provided by NIH BIOART^48^.

C-index performance showed a flat-to-slightly-negative relationship with event count (**Fig. S13-S14**). Across all conditions, Percival achieved the highest mean C-index in both anatomical regions (chest: 0.68; abdomen/pelvis: 0.70), exceeding TotalSegmentator (0.65, 0.66), CT-FM (0.61, 0.63), and the demographic baseline (0.61, 0.62; **Fig. 6a**). This pattern was consistent across all disease groups examined, with Percival outperforming each comparator in both regions (**Fig. S15**). Percival was successful at stratifying risk in 715 and 658 conditions for chest CT and abdominal/pelvis CT respectively. Restricting to chest CT, Percival’s C-index across conditions was broadly distributed within every disease group, indicating prognostic signal extending throughout the disease spectrum rather than concentrating in a few categories (**Fig. 6b**). Among respiratory conditions, Percival achieved a mean C-index of 0.70 (range=0.45, 0.95) across 76 conditions, including significant risk stratification for bronchiectasis (C-index=0.81; 95% CI=0.79, 0.84; **Fig. 6c**). Across circulatory conditions, Percival achieved a mean C-index of 0.72 across 127 conditions (range=0.56, 0.90), including heart failure with preserved ejection fraction (HFpEF; C-index=0.84; 95% CI=0.82, 0.85; **Fig. 6f**), a condition typically diagnosed via functional imaging. Amongst digestive conditions, Percival achieved a mean C-index of 0.64 across 129 conditions (range=0.45, 0.90), with significant prediction of cirrhosis (C-index=0.77; 95% CI=0.74, 0.81; **Fig. 6e**). When examining endocrine and metabolic conditions, Percival achieved a mean C-index of 0.69 across 94 conditions (range=0.49, 0.97), including strong prediction of cachexia (C-index=0.80; 95% CI=0.77, 0.83; **Fig. 6d**). Percival also stratified 116 genitourinary conditions (mean C-index=0.75, range=0.54, 0.91), highlighted by acute renal failure (C-index=0.72; 95% CI=0.71, 0.74; **Fig. 6h**), and 70 musculoskeletal conditions (mean C-index=0.64, range=0.47, 0.76), including osteoarthritis (C-index=0.69; 95% CI=0.66, 0.72; **Fig. 6g**). Additional prognostic signal was observed across neoplastic (83 conditions, mean C-index=0.71, range=0.56, 0.87) and hematopoietic conditions (37 conditions, mean C-index=0.67, range=0.46, 0.79). Complete chest CT results and the parallel abdomen/pelvis CT analysis are provided in **Tables S15-S16** respectively.

## DISCUSSION

This work introduces Percival, a large-scale CT vision-language model trained on more than 400,000 CT volumes from over 50,000 participants collected from multiple hospitals spanning the Penn Medicine Healthcare system. Prior work on CT vision-language modeling has primarily evaluated performance on task-specific benchmarks such as classification, retrieval, or report generation, often with supervised fine-tuning. In contrast, our work uses a heterogeneous, real-world biobank to characterize how a pretrained CT-VLM’s latent representations align with measured clinical, physiological, and longitudinal variables. These findings permit the following contributions: (1) we demonstrate that contrastive CT-report pretraining produces latent representations that align with demographic, physiological, and laboratory variation extending beyond imaging acquisition context and report content alone; (2) we systematically benchmark vision-language pretraining against alternative CT foundation-model paradigms across classification and prognostic modeling, representing the first comprehensive characterization of these paradigms at biobank scale and providing evidence that vision-language pretraining aligns with clinical information not fully accessible to vision-only or structured-anatomical approaches; and (3) we release Percival’s pretrained weights, training pipeline, and downstream model weights as an open-source resource, enabling external validation, reuse across biobanks, and continued investigation of medical foundation models. Together, these contributions provide a framework for using CT-VLMs as scientific instruments for studying multisystem health at biobank scale.

Recent CT vision-language models have established that contrastive pretraining on paired CT volumes and radiology reports is feasible at scale and yields representations that support a range of downstream tasks. Merlin demonstrated this for abdominal and pelvic CT, while CT-CLIP extended the approach to thoracic imaging^17,32^. Percival builds on these efforts by training across thoracic, abdominal, and pelvic CT volumes within a single shared representation space, enabling characterization of biological structure that extends across multiple anatomical fields of view. The CT-VLM literature, however, has largely evaluated representations through their performance on task-specific labels rather than through systematic characterization of what those representations encode about patient biology and longitudinal disease. Moreover, existing benchmarks have primarily compared CT-VLMs to other vision-language architectures or task-specific supervised baselines rather than to alternative foundation-model paradigms. This study extends the CT-VLM literature by quantifying the structure of a pretrained CT-VLM’s latent space across clinical, physiological, and longitudinal variation at biobank scale, and by benchmarking vision-language pretraining against alternative foundation-model paradigms.

Within the broader landscape of representation learning, contrastive vision-language pretraining sits alongside several alternative paradigms. Contrastive learning aligns paired image-text samples in a shared latent space while pushing unpaired samples apart, producing representations shaped jointly by both modalities that exceed what either modality conveys alone^33–35^. Vision-only contrastive learning, exemplified here by CT-FM, applies the contrastive objective to images alone (either intra-sample patch contrasts or inter-sample image contrasts), producing representations organized around visual concept identification without multimodal alignment via text^30^. Masked autoencoding takes a complementary route, learning representations through reconstruction of masked patches from visible patches^36^. Generative multimodal models adopt a distinct objective, producing representations through cross-modal generation such as image-conditional text generation^37^ or text-conditional image synthesis^38^, with representations shaped by the generative loss rather than by alignment. These paradigms differ in the kind of structure they impose on the latent space, and direct empirical comparison across all four at biobank scale remains an open direction for the field; in the present work, we benchmark against the two publicly available paradigms at comparable scale (vision-only contrastive, via CT-FM; structured anatomical features, via TotalSegmentator).

Using principal components derived from Percival’s latent feature space, we identified broad alignment between the latent representations and measured clinical and biological variables. Demographic and anthropometric signal was distributed across multiple principal components rather than concentrating on a single axis, with age and sex showing their strongest individual associations with PC2 and PC7 respectively. The laboratory-wide associations spanned multiple organ systems—renal, hepatic, metabolic, hematopoietic, and inflammatory—with 113 associations replicating across discovery and replication cohorts. These laboratory associations suggest that the latent space captures variation beyond imaging acquisition context or report content alone. When the same latent space was used to predict laboratory values directly, performance was weak in aggregate (mean R²=0.04) but substantially higher for a subset of clinically meaningful markers including albumin (R²=0.40), hemoglobin (R²=0.30), and eGFR (R²=0.26). These synthetic estimates do not replace clinical laboratory testing but may help identify individuals with extreme or unexpected biochemical values during opportunistic CT review^11,12^. We note that these associations should be interpreted as alignment with measured variables rather than as direct biological encoding by the model.

To examine the breadth and limits of how Percival’s latent representations align with temporal disease states, we evaluated associations between the latent feature space and diagnoses across the electronic health record. The latent space showed broad phenome-wide alignment with diagnoses spanning multiple disease groups, with associations replicating in a held-out cohort under stringent Bonferroni correction and adjustment for acquisition context. Although acquisition variables (region, contrast) constitute the dominant axes of variation in the latent space (**Fig. S3**), the phenome-wide associations persist after adjustment, indicating that disease-relevant signal lives beyond first-order acquisition clustering. Lifespan disease-burden mapping extended this analysis beyond cross-sectional association testing, revealing that participants in the extreme tails of specific latent components accumulated substantially elevated lifetime disease counts. For example, participants in the extreme tails of the 4th latent component accumulated more than 10 circulatory and more than 5 genitourinary diagnoses over longitudinal follow-up, indicating that the latent space tracks future multisystem morbidity rather than only conditions surrounding the imaging encounter.

To evaluate how CT-VLM latent representations compare to vision-only alternatives (CT-FM) and deterministic multi-organ segmentations (TotalSegmentator), we benchmarked Percival against both foundation-model paradigms in the disease classification setting. This comparison evaluates whether vision-language pretraining captures information beyond what vision-only contrastive learning or structured anatomical feature extraction provides. When examining structural diseases with clearly defined imaging correlates, Percival exhibited consistent performance gains relative to the alternative models. For coronary atherosclerosis, a disease with intensity-based imaging hallmarks such as vascular calcification, Percival outperformed TotalSegmentator on chest CT (AUC 0.85 vs 0.79). A similar pattern emerged for pancreatic cancer (AUC 0.91 vs 0.84), bronchiectasis, bacterial pneumonia, and cirrhosis, suggesting that Percival captures latent equivalents of the attenuation- and morphology-based features that drive these diagnoses. Percival also maintained strong classification performance for conditions whose primary clinical workup does not rely on CT, including atrial fibrillation, HFpEF, and chronic renal failure. Patients carrying these conditions often exhibit subclinical imaging changes (atrial fibrillation with left atrial enlargement; HFpEF with subtle chamber remodeling and pulmonary congestion; chronic renal failure with renal cortical thinning and atrophic kidneys) that may be present even when not the primary focus of clinical reporting. Contrastive image-text alignment surfaces these subclinical imaging variations as “colliding phenotypes” in the latent space, which are highly predictive of conditions whose primary diagnosis does not rely on CT. We interpret these findings as evidence that Percival has learned imaging features that co-occur with these diagnoses, not as evidence that the model has independently inferred their biology. Across both regions, Percival outperformed all four comparators on 279 conditions in chest CT and 230 conditions in abdomen/pelvis CT, with sex-stratified replication showing Percival’s cross-sex concordance matching TotalSegmentator on chest CT (R=0.85) and highest on abdomen/pelvis CT (R=0.83). Beyond discrimination, Percival also demonstrated the strongest calibration profile among the comparator models across both anatomical regions. Together, these results suggest that vision-language pretraining captures clinical information not fully accessible to vision-only contrastive learning or structured anatomical feature extraction alone.

In contrast to the classification setting, where performance scaled modestly with disease prevalence, prognostic C-index was not related to the number of incident events per condition; high-prevalence and low-prevalence outcomes were prognostically modeled with comparable performance. This decoupling suggests that the prognostic signal in Percival’s latent space is not primarily a function of how common a condition is, but reflects features tied to the longitudinal disease trajectory of the imaged participant. Across the prognostic benchmarks, Percival again outperformed comparison models. TotalSegmentator was the most competitive comparator in this setting, consistent with the prognostic value of organ volume and attenuation measurements, which on their own carry information about future disease risk. The vision-only contrastive paradigm (CT-FM), by contrast, lagged in prognostic discrimination despite remaining competitive in classification. A possible explanation lies in the structure of CT-FM’s pretraining objective: intra-sample contrastive learning contrasts patches from within a single scan against each other, producing representations organized around anatomical concept identification (heart, kidneys, lungs as distinct clusters) but with less explicit pressure to distinguish subtle within-organ variation. Vision-language pretraining, by contrast, aligns image embeddings with reports that routinely mention prognostically meaningful findings (e.g., coronary or aortic calcification preceding cardiovascular events; incidentally noted lung nodules preceding lung cancer), shaping the latent space around features that intra-sample contrastive objectives may not preferentially capture.

This study has notable strengths and some limitations. Percival was developed within a large clinical biobank of more than 70,000 participants with linked imaging and longitudinal health records. Over 50,000 participants contributed 400,000 CT volumes paired with radiology reports for training, spanning diverse fields of view, contrast phases, and acquisition settings. Validation on an additional 20,000 participants with more than 100,000 CT volumes enabled a robust assessment of generalizability across real-world imaging contexts. Notably, radiology reports are commonly associated with multiple CT volumes acquired in the same encounter; in such cases, radiologists summarize findings across all CT volumes in a single report, resulting in image-text pairs with incomplete mapping of features. Contrastive vision-language pretraining also relies on distinguishing paired image-text samples from unpaired ones; because most radiology reports describe normal, unremarkable, or negative findings, some negative image-text pairs may exhibit inherent similarity, and the model may learn to distinguish fine-grained variations in normal imaging patterns alongside pathological differences. Contrastive image-text pretraining shapes the latent space through both image content and the language radiologists use to describe it, and the relative contributions of these pathways to the observed phenotypic and prognostic associations cannot be fully disentangled. External validation through the CT-RATE benchmark established Percival’s performance on out-of-distribution thoracic classification tasks, although publicly available CT cohorts with linked laboratory and longitudinal diagnostic data were not available to externally replicate the broader biological, phenotypic, and prognostic analyses. The phenome-wide associations may also reflect index-event bias, where diagnoses surrounding the imaging encounter co-occur with the clinical indication for the scan.

Future extensions of CT-based vision-language models will benefit from deeper integration of biological information alongside imaging. Explicit incorporation of laboratory measurements, longitudinal clinical trajectories, and genomic or biomarker data could illuminate how CT-derived representations relate to measured biological variation across the electronic health record. Direct empirical comparison against alternative representation-learning paradigms at biobank scale, including masked autoencoding and generative multimodal models, would further clarify the contributions of vision-language alignment relative to other self-supervised objectives. Saliency and attention analyses, particularly when combined with TotalSegmentator organ segmentations, would also clarify which anatomical regions drive specific latent features, an important direction for follow-up work. We release Percival publicly to facilitate ongoing work in understanding and extending the biological and clinical capabilities of CT-based foundation models.

In summary, our study demonstrates that vision-language foundation models for three-dimensional CT imaging produce latent representations that align with measured clinical, physiological, and longitudinal variation extending beyond the findings typically associated with CT indications. To our knowledge, Percival is the most comprehensively trained foundation model for CT imaging to date, enabling a systematic characterization of how a CT-VLM’s latent space aligns with clinical, phenotypic, and prognostic variables. By releasing Percival publicly, we aim to provide a broadly accessible resource for advancing future work in precision medicine and for deepening our understanding of how medical foundation models relate to measured clinical and physiological variation.

## METHODS

### Study population

The Penn Medicine BioBank (PMBB) comprises more than 350,000 consenting patients who receive care in the Penn Medicine healthcare system^39^. Patients were enrolled during routine care appointments at a Penn Medicine location during which they provided written consent to either in-person or digitally to participate in the PMBB. This consent enabled the linkage of their electronic health records, including past disease diagnosis, laboratory measurements, and imaging data. PMBB imaging data were first de-identified by a third-party “honest broker,” who removed patient-identifying information from both the DICOM metadata and radiology reports (https://www.med.upenn.edu/radar/). For model training, we used 436,787 CT volumes paired with radiology reports from 51,966 PMBB participants. For validation, we included 123,603 CT volumes from 20,599 participants. The PMBB was approved by the Institutional Review Board at the University of Pennsylvania under IRB protocol 813913. The PMBB data is available under a data use agreement with the University of Pennsylvania, with access governed by HIPAA-compliant restrictions on re-identification, external sharing, and commercial use. This research was approved by the Institutional Review Board at the University of Pennsylvania under IRB protocol 857974.

### Image preprocessing

All CT volumes were acquired in a clinical setting using Siemens, GE HealthCare, Philips, Canon, or Hitachi scanners, stored as DICOM data directories, and subsequently converted to NIfTI format using the dcm2niix conversion tool^40^. All CT volumes were first reoriented to left posterior superior (LPS) coordinates and then resampled to a uniform voxel spacing of 1.0 x 1.0 x 3 mm (sagittal x coronal x axial), before being converted to Pytorch tensors to improve read times. Image radiodensities were clipped to a Hounsfield unit (HU) range of -1000 to 1000, followed by min-max normalization to a [0, 1] range. Finally, images were spatially padded and center-cropped to standardized dimensions of 352 x 352 pixels in-plane and 128 slices out-of-plane.

### VLM model architecture

Percival is composed of distinct vision and language encoders designed to establish latent feature space alignment between image and text modalities. We trained a family of Percival models spanning standard Vision Transformer scaling regimes (Tiny, Small, and Base), corresponding to approximately 6M, 22M, and 86M parameters in the vision encoder. All models were trained under a unified pipeline on the full PMBB corpus. Unless otherwise specified, all results in the main text refer to the Percival-Base configuration (∼86M parameters).

The vision encoder leverages a pretrained implementation of the Augmentation and Regularization Vision Transformers (AugReg) from the PyTorch Image Models (TIMM) library^41^. To process volumetric CT data, the vision transformer was adapted for three-dimensional imaging using three-dimensional patch embeddings, where each patch represents a 16 x 16 x 8 voxel segment that is projected into a latent embedding space using a 3D convolutional layer with kernel and stride equal to the patch size. From each CT volume, 7744 regional patch embeddings and a single global patch embedding are extracted to ensure comprehensive spatial representation. Within the transformer architecture, each regional patch embedding serves as a token input to the encoder layers, where local contextual relationships are modeled through multi-head self-attention (MSA). The global patch embedding functions analogously to a [CLS] token and participates in the same attention operations, attending to and being attended by all regional patches at each layer. Through this bidirectional exchange, the global embedding progressively integrates fine-grained anatomical and textural information from across the entire scan. The attention weights quantify the contribution of each regional patch to the global context, allowing the model to capture long-range dependencies between distant anatomical regions. For the language encoder, Percival employs the CXR-BERT-specialized architecture^42^. Image and text representations are aligned in a shared latent feature space whose dimensionality scales with the vision encoder configuration: 192, 384, and 768 dimensions for the Tiny, Small, and Base variants, respectively. A linear projection head maps both vision and text outputs to this shared space, where contrastive alignment is performed.

### Training protocol

Percival’s VLM pretraining used 436,787 paired three-dimensional CT volumes and radiology reports from 51,966 PMBB participants. Pretraining validation was performed on a held-out set of 46,535 paired CT volumes and radiology reports from 5,774 PMBB participants. An additional 77,068 CT volumes from 16,881 PMBB participants, for which paired radiology reports were not available, were included in the validation cohort for downstream analyses, which rely only on imaging data, bringing the combined validation cohort to 20,599 participants and 123,603 CT volumes. The vision and language encoders are jointly trained using a symmetric contrastive InfoNCE loss function, which optimizes the alignment between imaging and text representations^33,43,44^. This objective function maximizes cosine similarity between paired representations while minimizing similarity between unpaired representations, effectively distinguishing relevant cross-modal relationships. More specifically, given a batch of *b* image-text input pairs 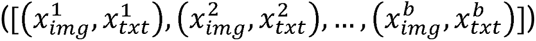, the vision and language encoders (*G_img_,G_txt_*) compute corresponding latent feature representations 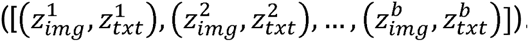. We then construct a similarity matrix *S* ɛ ℝ*^bxb^* where each element *S_i,j_* represents the cosine similarity between 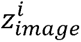 and 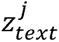. The diagonal of S contains the true paired image-text similarities, while the off-diagonal elements represent negative pairs. InfoNCE loss is subsequently computed using a temperature-scaled softmax:

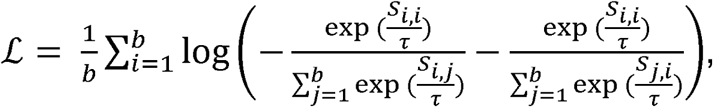

where τ is a temperature hyperparameter that controls the sharpness of the similarity distribution. This loss encourages image-text pairs to have high similarity while optimizer^45,46^ with a batch size of 48, τ = 10, and an initial learning rate of 5x10^-5^. The simultaneously pushing negative pairs apart. Training was performed using the AdamW Tiny, Small, and Base models were each trained on four NVIDIA B200 GPUs with PyTorch distributed data parallel (DDP), using a per-GPU batch size of 48; image and text features were gathered across GPUs prior to computing the contrastive loss, yielding an effective global batch size of 192 per training step.

### External validation: zero-shot classification and linear probing

To assess out-of-domain generalization, we evaluated Percival on the CT-RATE external validation cohort (n=1,564 CT volumes, with one CT volume retained per study), across 18 distinct thoracic pathology labels^17^. Percival was benchmarked against CT-CLIP, which was trained directly on the CT-RATE training data and therefore serves as an upper-bound reference for in-domain performance^17^. Two evaluation protocols were performed, namely (1) zero-shot classification and (2) linear probing. For zero-shot classification, the text encoder was provided with two standardized prompts: “[pathology] is present” and “[pathology] is not present.” The resulting text feature embeddings were compared to image embeddings using cosine similarity, and the prompt yielding the higher similarity score was selected as the predicted disease label. For linear probing, latent feature vectors were extracted from the CT-RATE training split by passing each volume through Percival’s frozen visual encoder; logistic regression classifiers were then trained on these embeddings to predict each thoracic pathology label and evaluated on the CT-RATE validation cohort.

### Comparator models for downstream analyses

To contextualize Percival’s downstream performance against alternative imaging-derived feature representations, we evaluated two publicly available comparator models on the PMBB validation cohort^13,30^. The first comparator was CT-FM, a vision-only three-dimensional CT foundation model pretrained on 148,000 CT volumes from the Imaging Data Commons using image-only contrastive learning^30^. CT-FM provides a matched contrastive baseline that differs from Percival primarily in the absence of paired text supervision, allowing the contribution of language alignment to be isolated. CT-FM embeddings were extracted by passing each CT volume through the vision encoder, yielding a 512-dimension feature vector per scan. The second comparator was TotalSegmentator, a deterministic 3D segmentation model that produces organ-level segmentation masks for thoracic, abdominal, and pelvic anatomical structures visible within each CT volume’s field of view^13^. From these segmentations, we extracted two scan-level features per structure (segmented volume in mL and mean radiodensity in HU). For the TotalSegmentator features, organ-level features missing for more than 50% of participants within a given anatomical field of view were excluded and remaining missing values were imputed using the training-set column mean. Comparator features were extracted on the same CT volumes used for Percival validation analyses and applied as predictors in the disease classification and prognostic risk modeling sections.

### t-SNE dimensionality reduction

To interrogate the unsupervised clustering structure of Percival’s latent feature space, we applied t-distributed stochastic neighbor embedding (t-SNE) for dimensionality reduction, followed by color-coding based on imaging field of view, contrast method, anatomical region, scanner manufacturer, age, sex, and BMI to assess unsupervised clustering patterns across both technical and demographic axes. Image latent features were extracted by passing the one CT volume per imaging accession from the training and validation sets through the vision encoder, generating a corresponding latent feature vector for each accession. These vectors were then projected into a two-dimensional space using t-SNE, allowing us to visually assess how the model organizes imaging data based on latent feature similarities.

### Association analyses of Percival’s latent space

To assess the biological and clinical relevance of Percival’s latent space, we performed principal component analysis (PCA) using one CT volume per imaging accession. First, we fit PCA on training-set embeddings. We then projected validation-set embeddings onto the train-fit components, ensuring that no validation-set variance contributed to the principal axes used in downstream association analyses. We restricted our association analyses to the top 10 components (PCs). Training-set embeddings were used solely to fit the PCA, and all subsequent association analyses were performed exclusively on the validation cohort. We focused on these leading components to provide a low-dimensional, interpretable summary of dominant axes of variation in the learned representation space and to enable stable statistical analysis across a wide range of clinical and laboratory traits. While additional variation is captured by higher-order components, our analysis focused on the principal modes of variation as broad statistical axes associated with clinical and demographic traits, rather than as an exhaustive characterization of the latent space. For each PC we evaluated associations with nine continuous physiological traits: age at imaging, height (cm), weight (kg), body mass index (BMI), systolic and diastolic blood pressure (mmHg), pulse (bpm), oxygen saturation (%), and respiratory rate (breaths/min). Each PC-trait pair was modeled both as a univariate linear regression and as a covariate-adjusted regression that controlled for age, sex, anatomical region, contrast type, and the imaging-to-measurement time offset (with the trait being modeled excluded from its own adjuster set). For each trait, only measurements obtained within a per-trait tolerance window of imaging were retained (±365 days for height and weight; ±90 days for blood pressure, pulse, oxygen saturation, and respiratory rate). Participants with missing values or measurements outside the imaging-to-measurement window were excluded from that trait’s association analysis. When multiple values fell within the window, the closest-in-time value was retained. Sex differences within each PC were quantified using analysis of variance with Hedges-corrected Cohen’s D as the effect-size measure. Significance was determined following Bonferroni correction across PC x trait tests for the continuous block (*P*<0.05/90) and across PCs for the sex block (*P*<0.05/10). We then conducted a laboratory-wide association study (LabWAS) using a half-half discovery-replication design to evaluate whether these components captured biologically meaningful variation. For each of the 64 laboratory traits in the PMBB validation cohort, we fit separate linear regression models for each of the 10 PCs, adjusting for age, sex, anatomical region, contrast type, and the time interval between imaging and laboratory or clinical measurement. Only laboratory values obtained within 60 days of imaging were included in the analysis. Significance of associations was determined by Bonferroni correction (*P*<0.05/640) in both the discovery and replication cohorts, with directional concordance of effect estimates required between cohorts.

### Synthetic laboratory value estimation

For each laboratory measurement, we developed synthetic laboratory estimation models using linear probing of Percival’s latent feature space in the PMBB validation cohort. Each model was implemented as a three-layer multilayer perceptron with hidden layer sizes of 256 and 128 units, ReLU activations, and a dropout rate of 0.2, trained with AdamW (learning rate 1x10^-3^, weight decay 1x10^-4^) using mean-squared-error loss. Predictor variables included Percival’s complete latent feature space as well as the difference between the date of imaging and the date of lab measurement. Models were trained and evaluated using five-fold subject-grouped cross-validation, so that no participant appeared in both the training and validation folds of any iteration. Model performance was quantified using the coefficient of determination (R^2^), Pearson correlation, and root mean squared error on the held-out folds. During inference, the number of days between imaging and assay is set to 0, thus enabling synthetic estimation at the time of imaging.

### Phenotypic associations

We performed a phenome-wide association study (PheWAS) to evaluate associations between Percival’s latent feature space and clinical disease states. Positive cases were defined using ICD-10CM codes recorded within 60 days of imaging in the PMBB validation cohort. ICD-10CM codes were mapped to phecodes using the PheWAS package. Where multiple ICD-10CM codes mapped to the same phecode, the code with the greatest number of positive cases was retained. For participants with repeat imaging studies each containing multiple CT volume acquisitions, one CT volume per imaging accession was retained. PheWAS modeling was performed using a half-half discovery-replication design: PCs derived from training-set embeddings were applied to the discovery and replication cohorts, and logistic regression was fit separately within each. Each model adjusted for age, sex, anatomical region, and contrast type. Phenome-wide significance was determined following Bonferroni correction (*P*<0.05/number of phecodes) in the discovery and replication cohort, with directional concordance between cohorts. To assess robustness to the temporal window between imaging and diagnosis, sensitivity analyses were conducted at progressively narrower windows of 30, 15, and 7 days.

### Lifespan Disease Burden Mapping

To assess global disease accumulation as a function of latent features, we quantified post-imaging disease burden for all participants in the validation cohort using ICD-10CM codes recorded after the date of imaging. ICD codes were mapped to phecodes using the PheWAS package and each phecodes was assigned to a phecodes group (e.g., circulatory, respiratory, endocrine/metabolic). Disease burden was summarized for each participant as the total number of unique phecodes and the number of unique phecodes within each group. Each principal component was z-score normalized and discretized into integer bins (-3 to +3). Average disease burden per-participant was computed within each bin and visualized as a heatmap to reveal global trends linking latent features to future disease accumulation.

### Percival classification models

To perform temporal disease-state classification, we developed LASSO-penalized logistic regression models for classifying disease states using Percival’s complete latent feature space as predictors. Input features were normalized prior to model fitting, and the L1 regularization strength was selected by 3-fold cross-validation on the training cohort. Analyses were stratified by anatomical field of view, with separate models trained for chest and abdomen/pelvis CT volumes. For participants with repeat imaging studies, one CT volume per imaging accession was retained. ICD-10CM codes were mapped to phecodes using the PheWAS package, and where multiple ICD-10CM codes mapped to the same phecode, the code with the greatest number of cases was retained. Codes occurring within 60 days of imaging were used to define positive cases. Conditions with fewer than 50 events in the training cohort or fewer than 20 events in the validation cohort were omitted from the analysis. Models were trained on the training cohort and evaluated on the held-out validation cohort using area under the receiver operating characteristic curve (AUC). In addition to AUC, model performance was assessed using Brier score, and calibration slope and intercept to characterize the calibration of predicted probabilities^47^. To evaluate the predictive value of the latent feature space, model performance was compared to two baseline models, the first including only age and sex, and the second additionally including the total number of prior imaging accessions to control for healthcare utilization, and to comparator models built from CT-FM embeddings and TotalSegmentator-derived organ features^13,30^. Statistical significance was assessed using the DeLong test, with FDR correction applied for multiple comparisons and significance established at the 5% threshold^31^. 95% confidence intervals for all reported metrics were computed by paired percentile bootstrap with 1,000 resamples of the validation cohort, with identical resample indices applied across comparator models.

### Percival prognostic risk modeling

To assess prognostic utility, we developed Cox proportional-hazards models for clinical disease states using Percival’s complete latent feature space as predictors, with ridge regularization. Analyses were stratified by anatomical field of view, with separate models trained for chest and abdomen/pelvis CT volumes. For participants with repeat imaging studies, the earliest CT volume in the participant’s electronic health record was retained. Time-to-event outcomes were defined using ICD-10CM codes, with participants censored at their most recent encounter in the electronic health record. ICD-10CM codes were mapped to phecodes using the PheWAS package, and where multiple ICD-10CM codes mapped to the same phecode, the code with the greatest number of cases was retained. Participants with a recorded diagnosis on or before the date of imaging were excluded from the corresponding condition’s analysis. Models were trained on the training cohort and evaluated on the held-out validation cohort using the concordance index (C-index). C-index confidence intervals (95%) were computed by paired percentile bootstrap (1,000 resamples of the validation cohort with models held fixed, identical resample indices applied across all comparator models). Conditions with fewer than 50 events in the training cohort or fewer than 20 events in the validation cohort were omitted from the analysis. Percival’s performance was compared against a baseline containing only age and sex, as well as against comparator models built from CT-FM embeddings and TotalSegmentator-derived organ features^13,30^. For visualization of risk stratification, participants were divided into low-risk (bottom 80%) and high-risk (top 20%) groups based on the linear predictor from the fitted Cox model, with group differences in event-free survival assessed using the log-rank test.

## Supporting information

Supplemental Note: PMBB leadership

## AUTHOR CONTRIBUTIONS

C.A.B. carried out model and code development, designed experiments, and drafted the manuscript.

W.R.W. supervised the project, assisted with experiment design and code development, acquired funding, and drafted and reviewed the manuscript.

J.K., B.H., R.S.1, R.S.2, E.E., G.Z., T.C., and J.D.1 contributed to code development and/or experiments.

J.D.2, A.V., and C.M. processed and curated clinical imaging, text, EHR, ICD, and laboratory data.

F.D., H.S., S.D., R.S.2, C.E.K., and J.A.C. provided clinical input to experiment design and interpretation of results.

H.T., B.Z., Q.L., L.S., and C.D. contributed to experiment design, statistical analysis/interpretation, or computational infrastructure and revised the manuscript.

J.G., M.D.R. and D.R. contributed to study conception and interpretation of results and provided financial support.

PMBB refers to the Penn Medicine Biobank. A full list of contributions from the Penn Medicine Biobank team is provided in supplement.

All authors reviewed the manuscript

## DATA AVAILABILITY

Access to Penn Medicine BioBank data is provided to investigators at the University of Pennsylvania. Percival’s model architecture, pretrained weights, classification and prognostic models have been made available: https://github.com/cams2b/percival

## ACKNOWLEDGEMENTS

We acknowledge the Penn Medicine BioBank (PMBB) for providing data and thank the patient-participants of Penn Medicine who consented to participate in this research program. We would also like to thank the Penn Medicine BioBank team and Regeneron Genetics Center for providing genetic variant data for analysis. The PMBB is approved under IRB protocol# 813913 and supported by Perelman School of Medicine at University of Pennsylvania, a gift from the Smilow family, and the National Center for Advancing Translational Sciences of the National Institutes of Health under CTSA award number UL1TR001878. Research reported in this publication was supported by the National Heart, Lung, And Blood Institute of the National Institutes of Health under Award Number F31HL182332. The content is solely the responsibility of the authors and does not necessarily represent the official views of the National Institutes of Health.

## FUNDING

C.A.B is supported by NIH grant F31-HL182332. W.R.W. is supported by NIH grants P41-EB029460, R01-HL169378, R01-HL137984, UL1-TR001878, R21-EB036734, OT2-OD038048. J.A.C. is supported by NIH grants R01-HL121510, R33-HL146390, R01-HL153646, R01-AG058969, R01-HL104106, P01-HL094307, R03-HL146874, and K24-AG070459. J.G is supported by R01-EB031722, R01-HL133889. L.S. is supported by P30-AG073105, U01-AG088658 and U01-AG066833. MDR is supported by UL1-TR001878. E.E is supported by NIH grant OT2-OD038048.

## DISCLOSURE

Dr. Chirinos is supported by NIH grants U01-TR003734, U01-TR003734-01S1, UO1-HL160277, R33-HL146390, R01-HL153646, K24-AG070459, R01-AG058969, R01-HL157108, R01-HL155599, R01-HL104106 and R01HL155764. He has recently consulted for Bayer, Fukuda-Denshi, Bristol-Myers Squibb, Biohaven Pharmaceuticals, Johnson & Johnson, Edwards Life Sciences, Merck, and NGM Biopharmaceuticals. He received University of Pennsylvania research grants from National Institutes of Health, Fukuda-Denshi, Bristol-Myers Squibb, Microsoft and Abbott. He is named as inventor in a University of Pennsylvania patent for the use of inorganic nitrates/nitrites for the treatment of Heart Failure and Preserved Ejection Fraction and for the use of biomarkers in heart failure with preserved ejection fraction. He has received payments for editorial roles from the American Heart Association, the American College of Cardiology, Elsevier and Wiley, and payments for academic roles from the University of Texas, Boston University, and Virginia Commonwealth University. He has received research device loans from Atcor Medical, Fukuda-Denshi, Unex, Uscom, NDD Medical Technologies, Microsoft and MicroVision Medical. The remaining authors have nothing to disclose.

## Notes

### Competing Interest Statement

Dr. Chirinos is supported by NIH grants U01-TR003734, U01-TR003734-01S1, UO1-HL160277, R33-HL-146390, R01-HL153646, K24-AG070459, R01-AG058969, R01-HL157108, R01-HL155599, R01-HL104106 and R01HL155764. He has recently consulted for Bayer, Fukuda-Denshi, Bristol-Myers Squibb, Biohaven Pharmaceuticals, Johnson & Johnson, Edwards Life Sciences, Merck, and NGM Biopharmaceuticals. He received University of Pennsylvania research grants from National Institutes of Health, Fukuda-Denshi, Bristol-Myers Squibb, Microsoft and Abbott. He is named as inventor in a University of Pennsylvania patent for the use of inorganic nitrates/nitrites for the treatment of Heart Failure and Preserved Ejection Fraction and for the use of biomarkers in heart failure with preserved ejection fraction. He has received payments for editorial roles from the American Heart Association, the American College of Cardiology, Elsevier and Wiley, and payments for academic roles from the University of Texas, Boston University, and Virginia Commonwealth University. He has received research device loans from Atcor Medical, Fukuda-Denshi, Unex, Uscom, NDD Medical Technologies, Microsoft and MicroVision Medical. The remaining authors have nothing to disclose.

### Author Declarations

The PMBB was approved by the Institutional Review Board at the University of Pennsylvania under IRB protocol 813913. This research was approved by the Institutional Review Board at the University of Pennsylvania under IRB protocol 857974.

### Summary of Updates

This version of the manuscript has been revised to include additional model architecture details as well as comparisons with alternative CT models.

