## Supplemental Figures for "Generalizable CT Vision-Language Modeling for Population Health and Disease Risk"

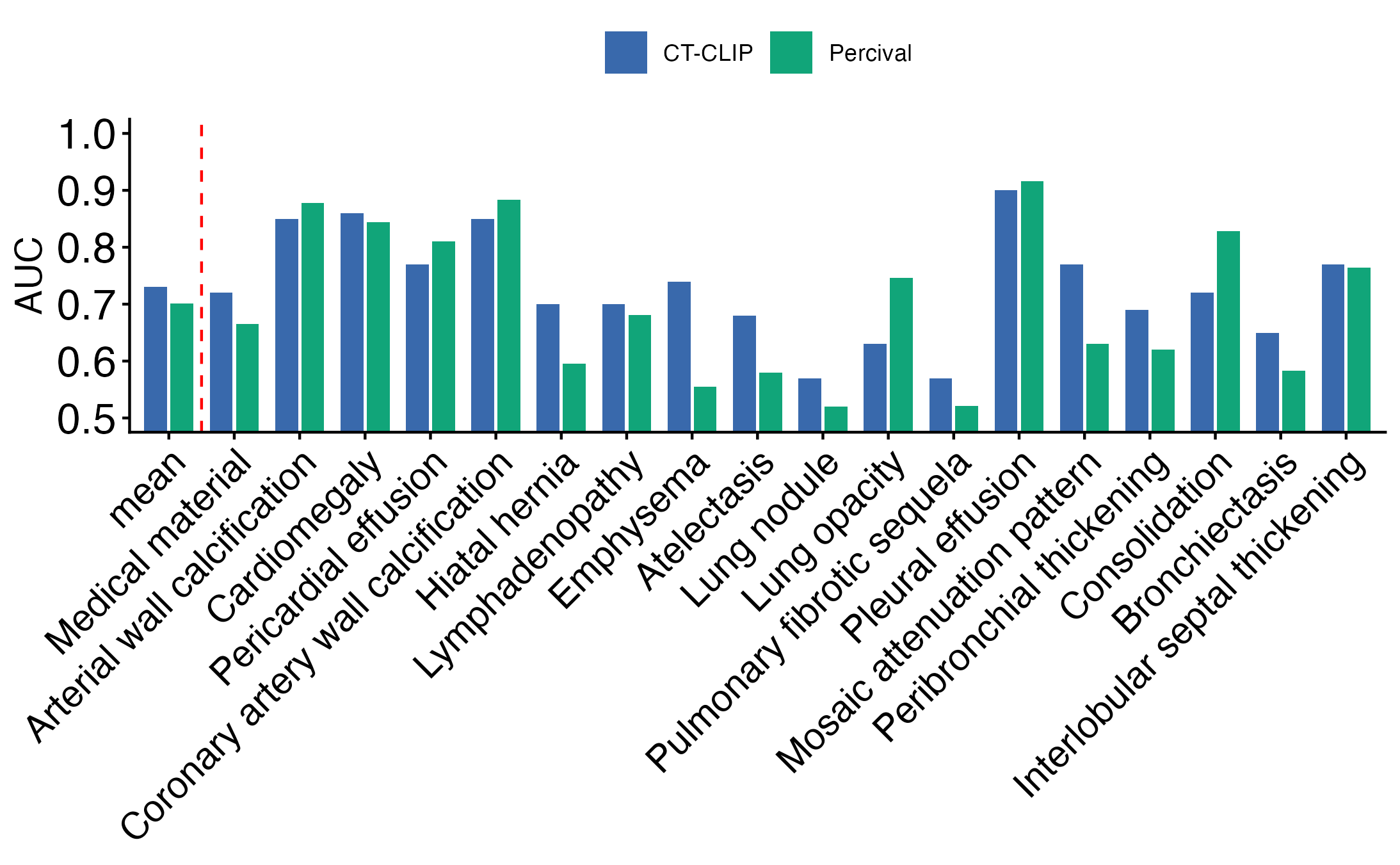

**Figure S1.** Zero-shot classification performance of Percival and CT-CLIP on the CT-RATE external validation cohort (n = 1,564) across 18 thoracic pathology labels. Bars show area under the receiver operating characteristic curve (AUC) per pathology, with the leftmost group ("mean") representing the average across all 18 labels. CT-CLIP is shown for reference as the in-domain upper bound, having been trained on the CT-RATE training split. Percival was evaluated without any fine-tuning or exposure to CT-RATE during training. CT-CLIP performance values are from the original publication^1^.

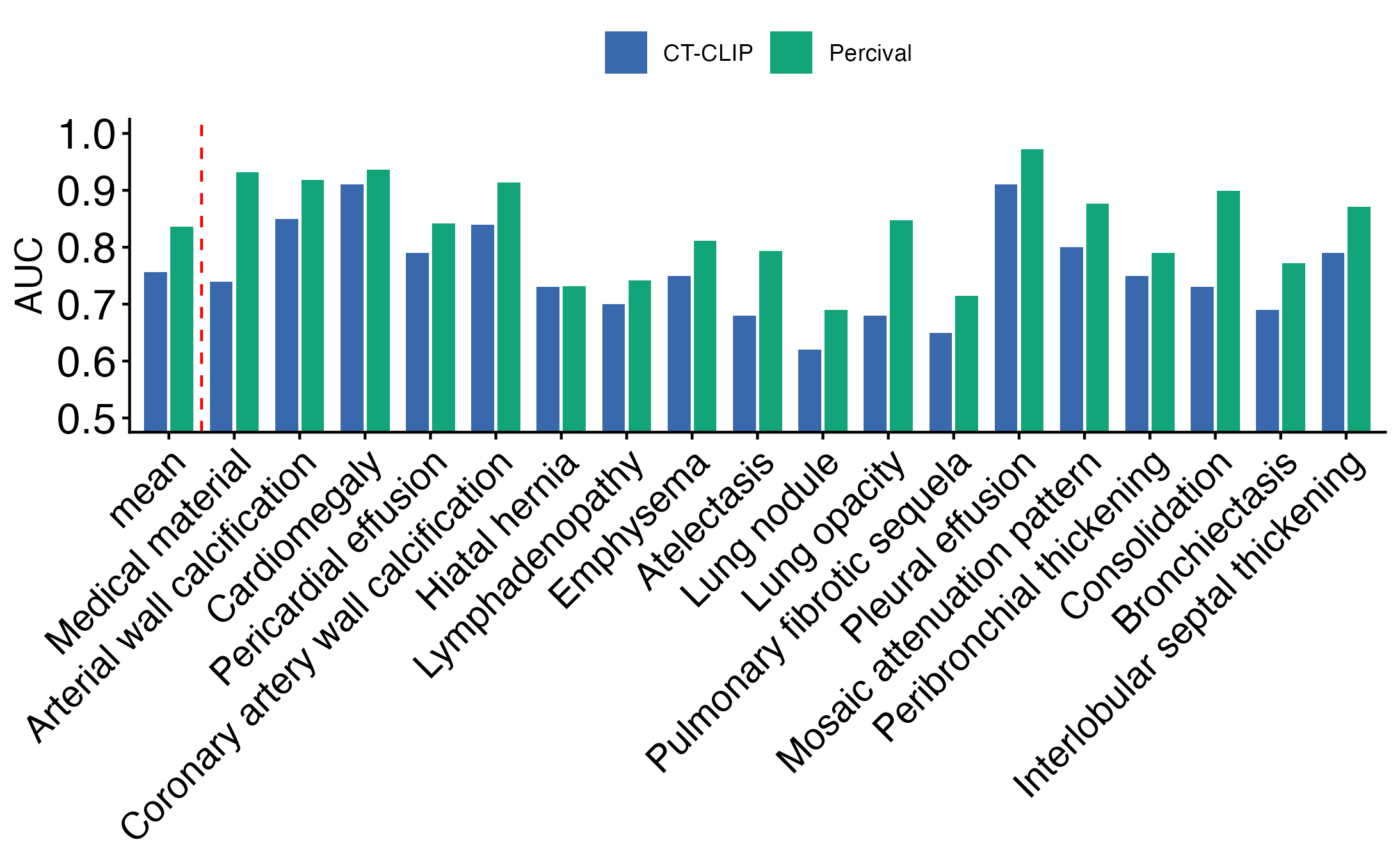

**Figure S2.** Linear probing performance of Percival and CT-CLIP on the CT-RATE external validation cohort (n = 1,564) across 18 thoracic pathology labels. Bars show AUC per pathology, with the leftmost group ("mean") representing the average across all 18 labels. CT-CLIP is shown for reference as the in-domain upper bound, having been trained on the CT-RATE training split. Linear classifiers were trained and evaluated on Percival or CT-CLIP embeddings extracted from the CT-RATE validation cohort. CT-CLIP performance values are from the original publication^1^.

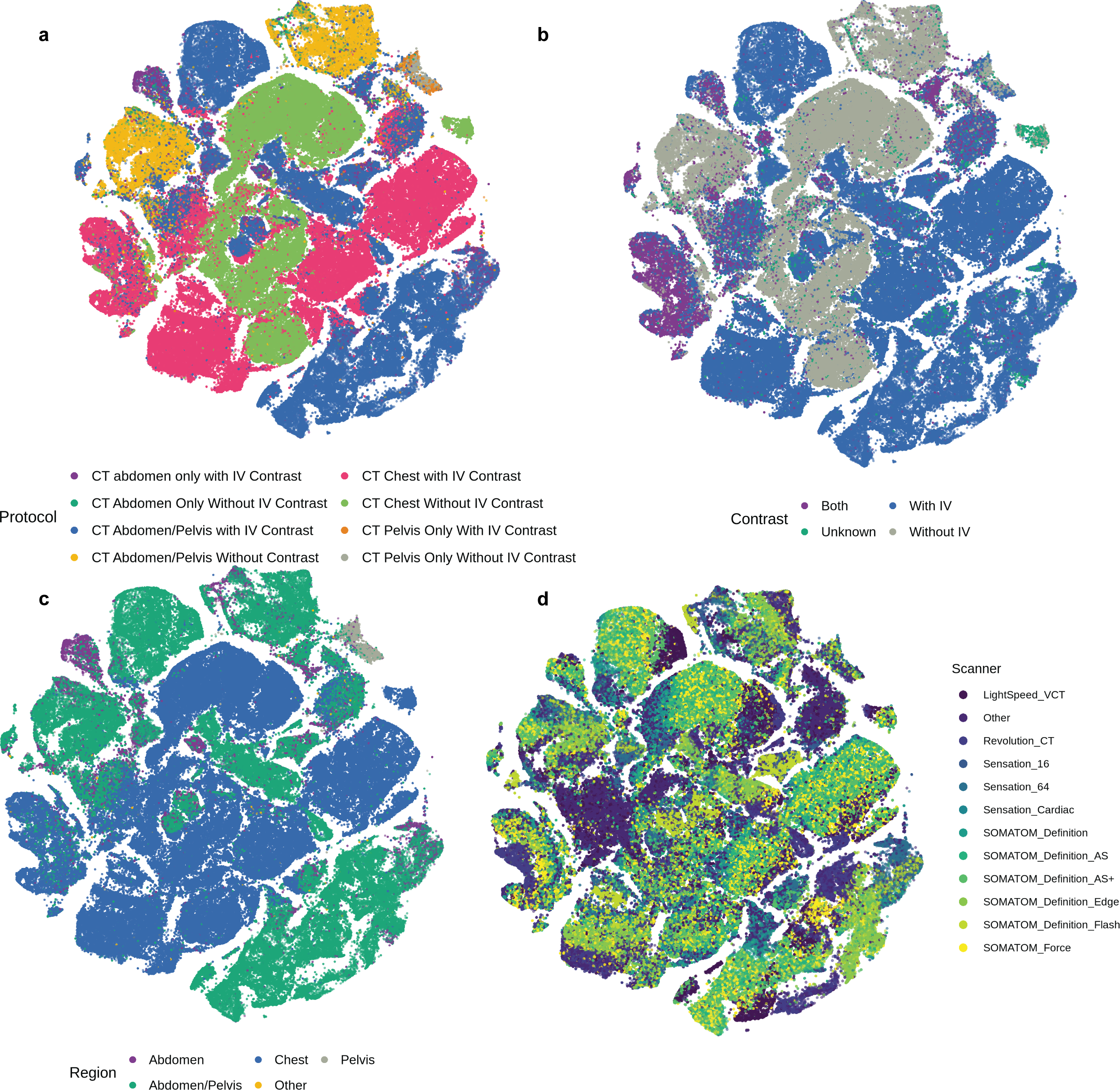

**Figure S3.** Two-dimensional t-distributed stochastic neighbor embedding (t-SNE) of Percival's 768-dimensional latent feature space, computed across the complete PMBB cohort, with each point representing one imaging accession colored by imaging acquisition attributes. (**a**) Combined imaging protocol (field of view × intravenous contrast), spanning eight categories from CT abdomen-only and pelvis-only through abdomen/pelvis and chest acquisitions with and without IV contrast. (**b**) Intravenous contrast administration (with IV contrast, without IV contrast, both, or unknown). (**c**) Anatomical region (chest, abdomen, pelvis, abdomen/pelvis, other). (**d**) Scanner manufacturer; manufacturer identity is intermixed throughout the embedding rather than forming distinct clusters, indicating that scanner identity is not a dominant axis of variation in Percival's latent space. Strong spatial separation is observed for imaging protocol, contrast administration, and anatomical region, while scanner manufacturer does not produce distinct clustering.

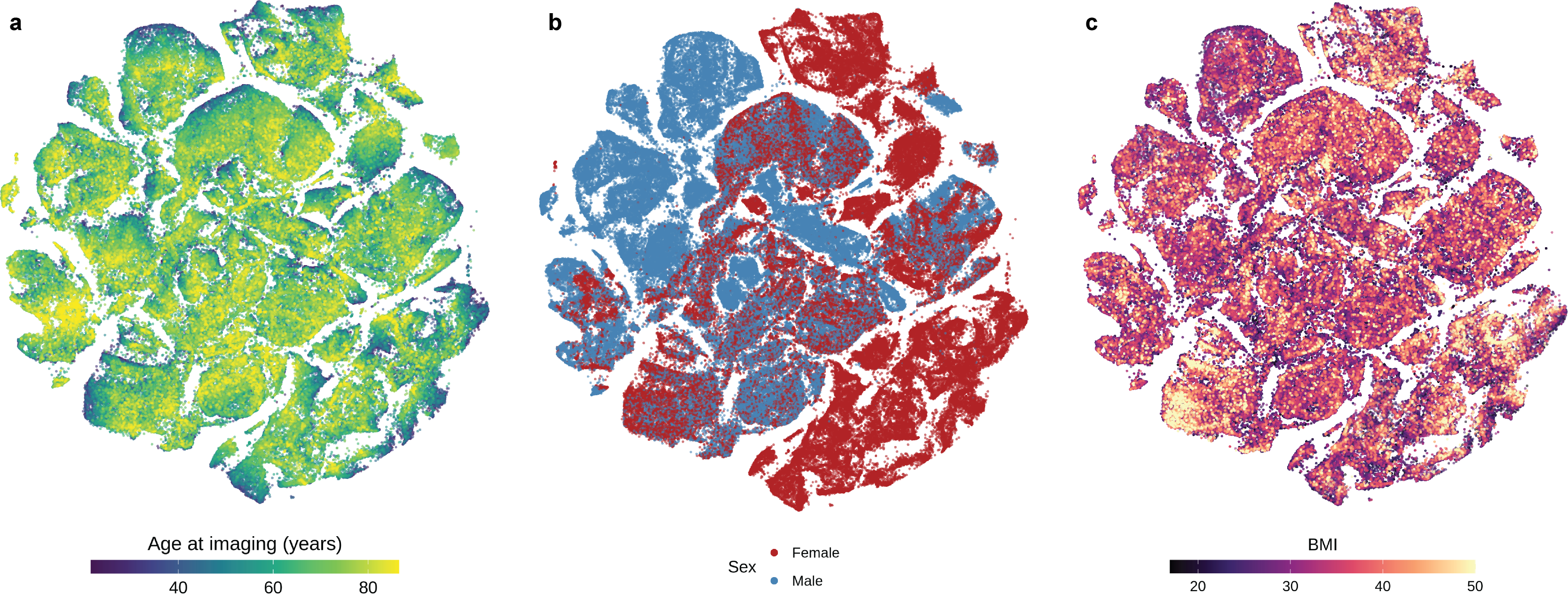

**Figure S4.** Two-dimensional t-distributed stochastic neighbor embedding (t-SNE) of Percival's 768-dimensional latent feature space, computed across the complete PMBB cohort, with each point representing one imaging accession colored by participant demographic attributes. (**a**) Age at imaging (years). (**b**) Sex (male, female). (**c**) Body mass index (BMI). Distinct organization across the embedding is observed for each attribute, with sex (**b**) producing the clearest spatial separation, age (**a**) showing a graded distribution across the embedding, and BMI (**c**) showing localized regions of higher-BMI concentration. Together, these patterns indicate that Percival's latent space captures demographic variation alongside imaging features.

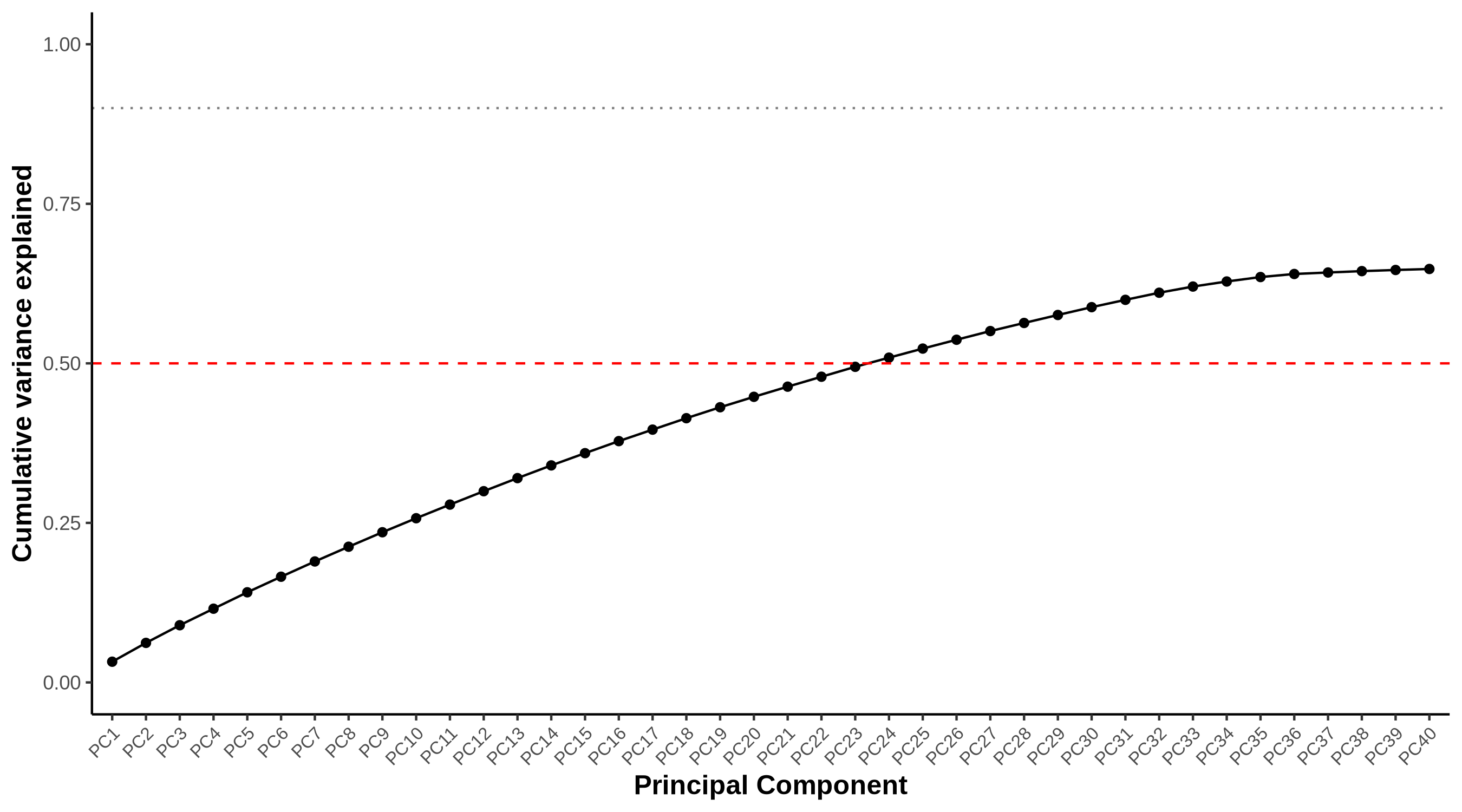

**Figure S5.** Cumulative variance explained by the first 40 principal components derived from Percival’s latent feature embeddings. The x-axis represents individual principal components (PCs), and the y-axis indicates the cumulative proportion of variance explained. A dashed red line marks the 50% threshold.

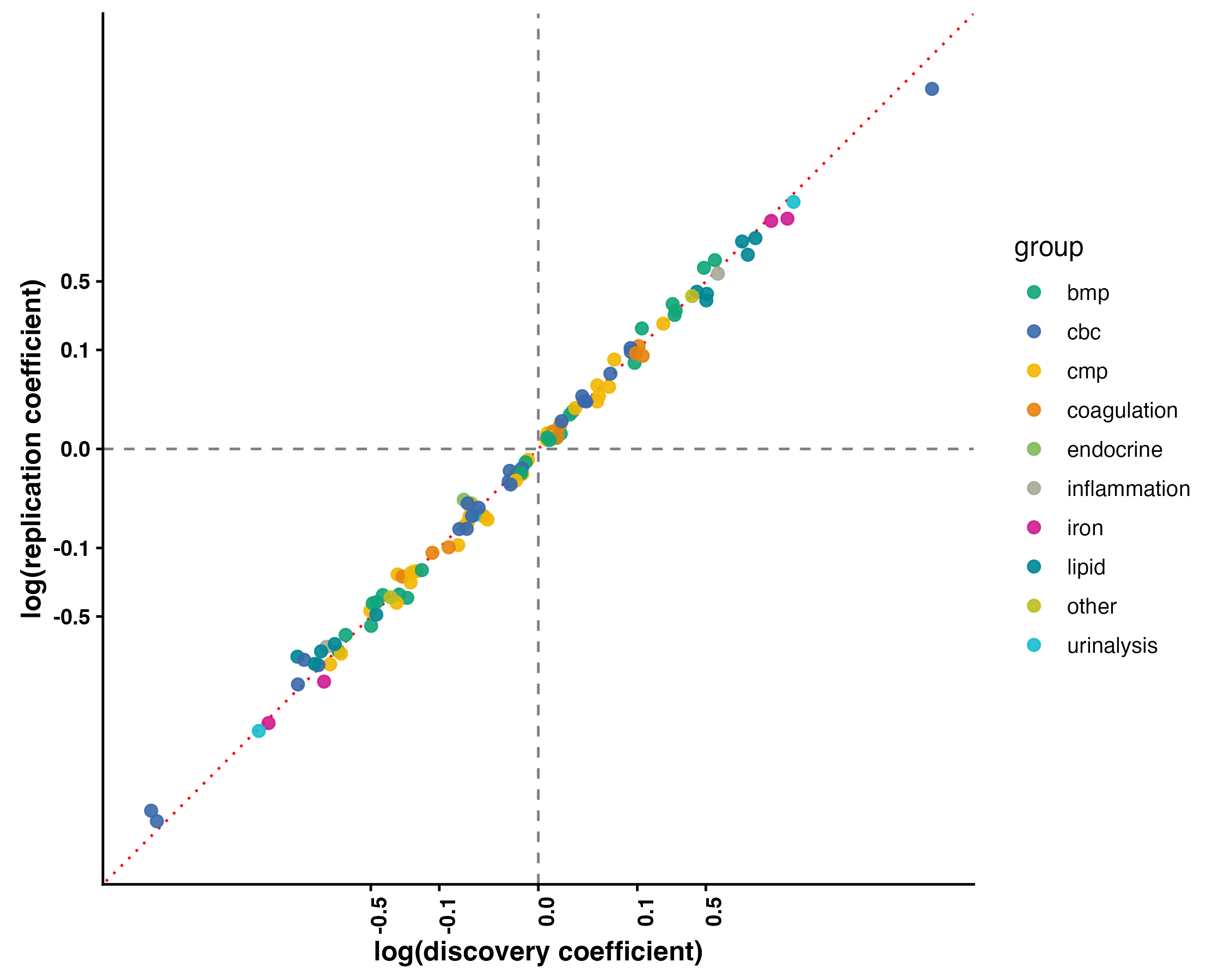

**Figure S6.** Discovery-replication concordance of significant associations between Percival's latent feature space and clinical laboratory measurements. Each point represents one principal component × laboratory measurement association that reached Bonferroni significance (*P*<0.05/640) in both the discovery (average n=2,652) and replication (average n=2,606) cohorts and shared effect direction across cohorts. The x- and y-axes show the log-transformed regression coefficients from the discovery and replication cohorts, respectively. Points are colored by laboratory group (basic metabolic panel, complete blood count, comprehensive metabolic panel, coagulation, endocrine, inflammation, iron, lipid, urinalysis, other). The diagonal red dotted line indicates perfect concordance; the dashed lines mark zero.

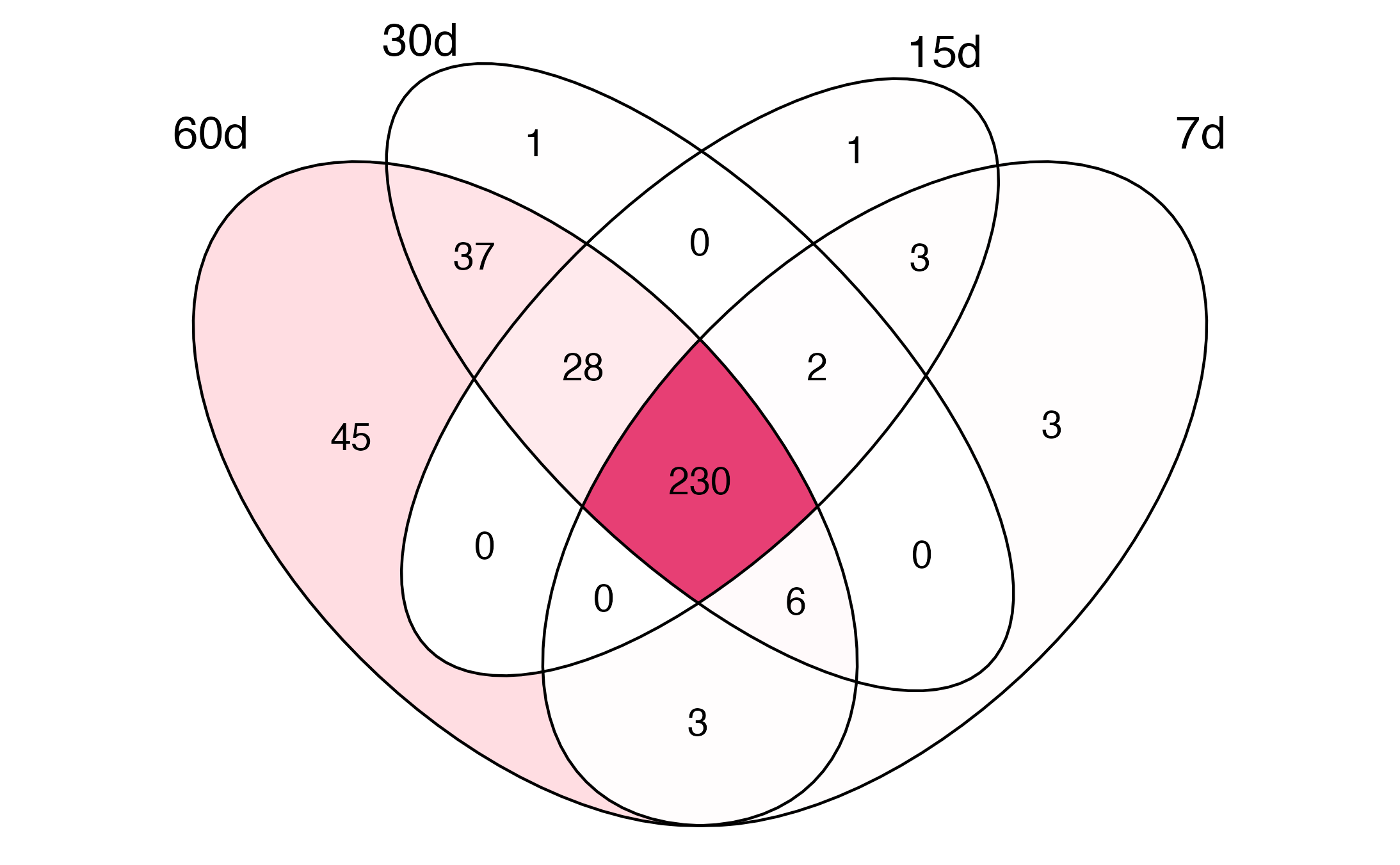

**Figure S7.** Concordance of PheWAS associations across imaging-to-diagnosis temporal windows. Each ellipse represents the set of phecodes reaching Bonferroni-significant association with Percival's principal components (PCs) in both the discovery and replication cohorts at one temporal window (60, 30, 15, or 7 days between imaging and diagnosis). Counts indicate the number of phecodes shared across each intersection. A total of 230 phecodes (center) reached phenome-wide significance across all four windows, indicating that the associations reported in the main analysis are robust to narrowing of the temporal proximity threshold.

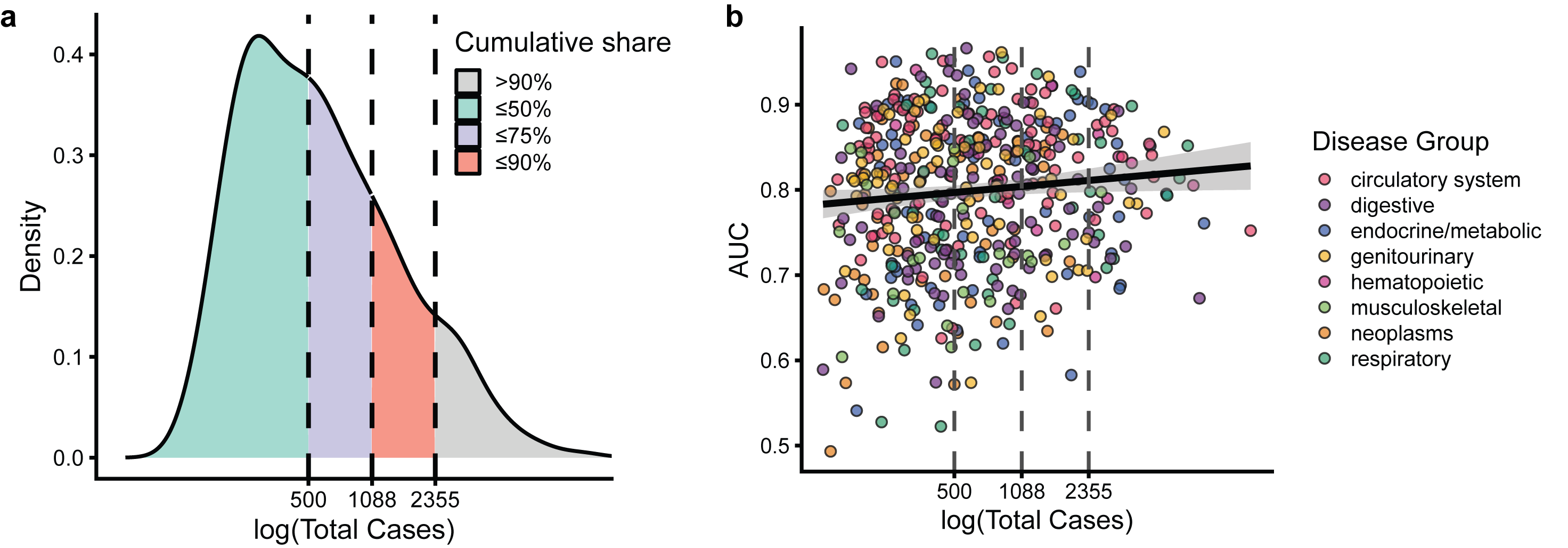

**Figure S8.** Distribution of positive case counts and discriminative performance across abdomen/pelvis disease classification models. (**a**) Density distribution of the number of positive cases per condition across 899 phecodes, shaded by cumulative share quartiles. Dashed vertical lines mark the 50th (n=500), 75th (n=1,088), and 90th (n=2,355) percentiles of the case-count distribution. (**b**) Scatter plot of Percival's AUC against the log-transformed number of positive cases per condition, with points colored for select phecode groups. The fitted line and shaded band show a linear regression with 95% confidence interval. Vertical dashed lines mark the same percentile thresholds as in panel (**a**).

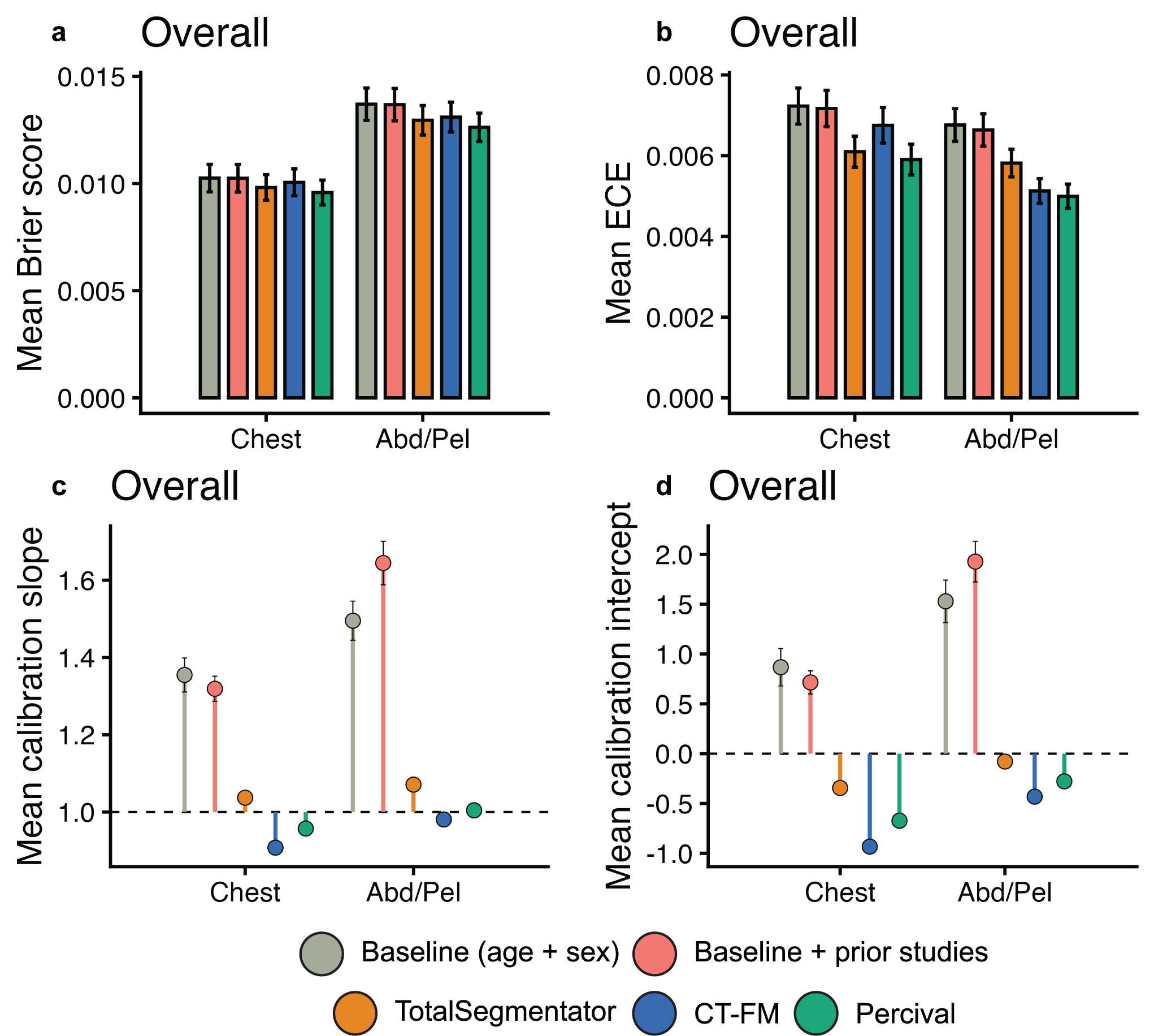

**Figure S9.** Overall calibration performance for (**1**) baseline (age + sex), (**2**) baseline + prior studies (referring to all previous studies accumulated up to the point of imaging), (**3**) CT-FM, a vision-only contrastive learning foundation model, (**4**) TotalSegmentator, a deterministic organ-level segmentation model, and (**5**) Percival. Calibration is summarized across all disease classification tasks for chest and abdomen/pelvis (Abd/Pel) CT using four metrics: mean Brier score (**a**), mean expected calibration error (ECE; **b**), mean calibration slope (**c**), and mean calibration intercept (**d**). For Brier score and ECE, lower values indicate better-calibrated probabilities. For calibration slope and intercept, dashed reference lines denote perfect calibration (slope = 1.0, intercept = 0.0).

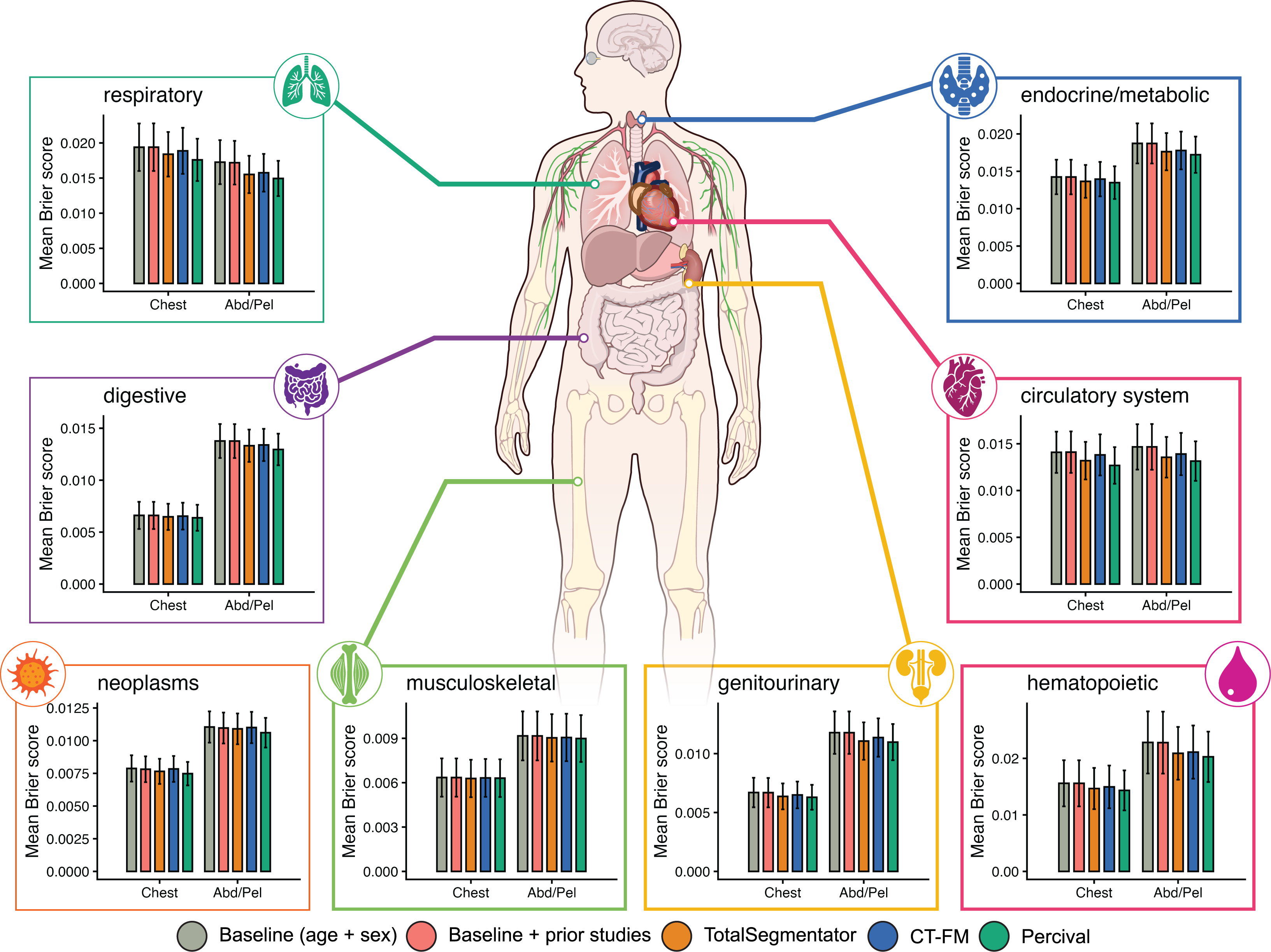

**Figure S10.** Disease-group-specific calibration performance for (**1**) baseline (age + sex), (**2**) baseline + prior studies (referring to all previous studies accumulated up to the point of imaging), (**3**) CT-FM, a vision-only contrastive learning foundation model, (**4**) TotalSegmentator, a deterministic organ-level segmentation model, and (**5**) Percival. Mean Brier score is shown across disease classification tasks within each of eight disease groups (respiratory, endocrine/metabolic, digestive, circulatory system, neoplasms, musculoskeletal, genitourinary, and hematopoietic), separately for chest and abdomen/pelvis (Abd/Pel) CT. Lower Brier scores indicate better-calibrated probability estimates. Error bars represent the standard deviation of Brier scores across conditions within each disease group. Human anatomical art provided by NIH BIOART^2^.

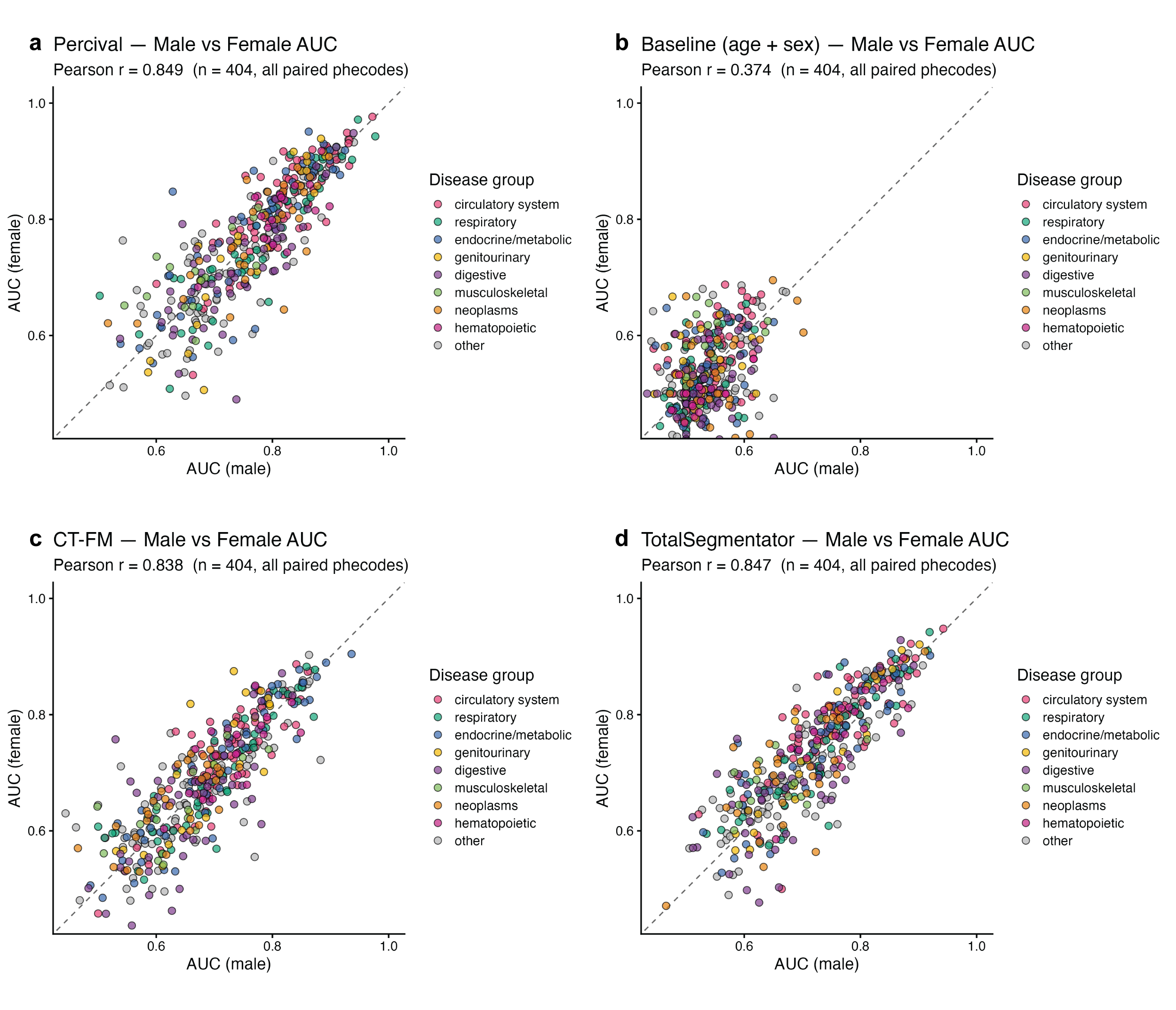

**Figure S11.** Sex-specific replication of disease classification performance in chest CT, comparing Percival to the demographic baseline, CT-FM, and TotalSegmentator. Each point represents one phecode plotted by area under the receiver operating characteristic curve (AUC) computed separately in male and female participants (n=404 phecodes). Points are colored by disease group, with phecodes outside the eight major groups shown as "other." The dashed diagonal indicates perfect cross-sex concordance. Pearson r is annotated in each panel. (**a**) Percival shows the highest cross-sex concordance (Pearson r=0.849). (**b**) Demographic baseline of age and sex shows the lowest concordance (Pearson r=0.374). (**c**) CT-FM (Pearson r=0.838). (**d**) TotalSegmentator (Pearson r=0.847).

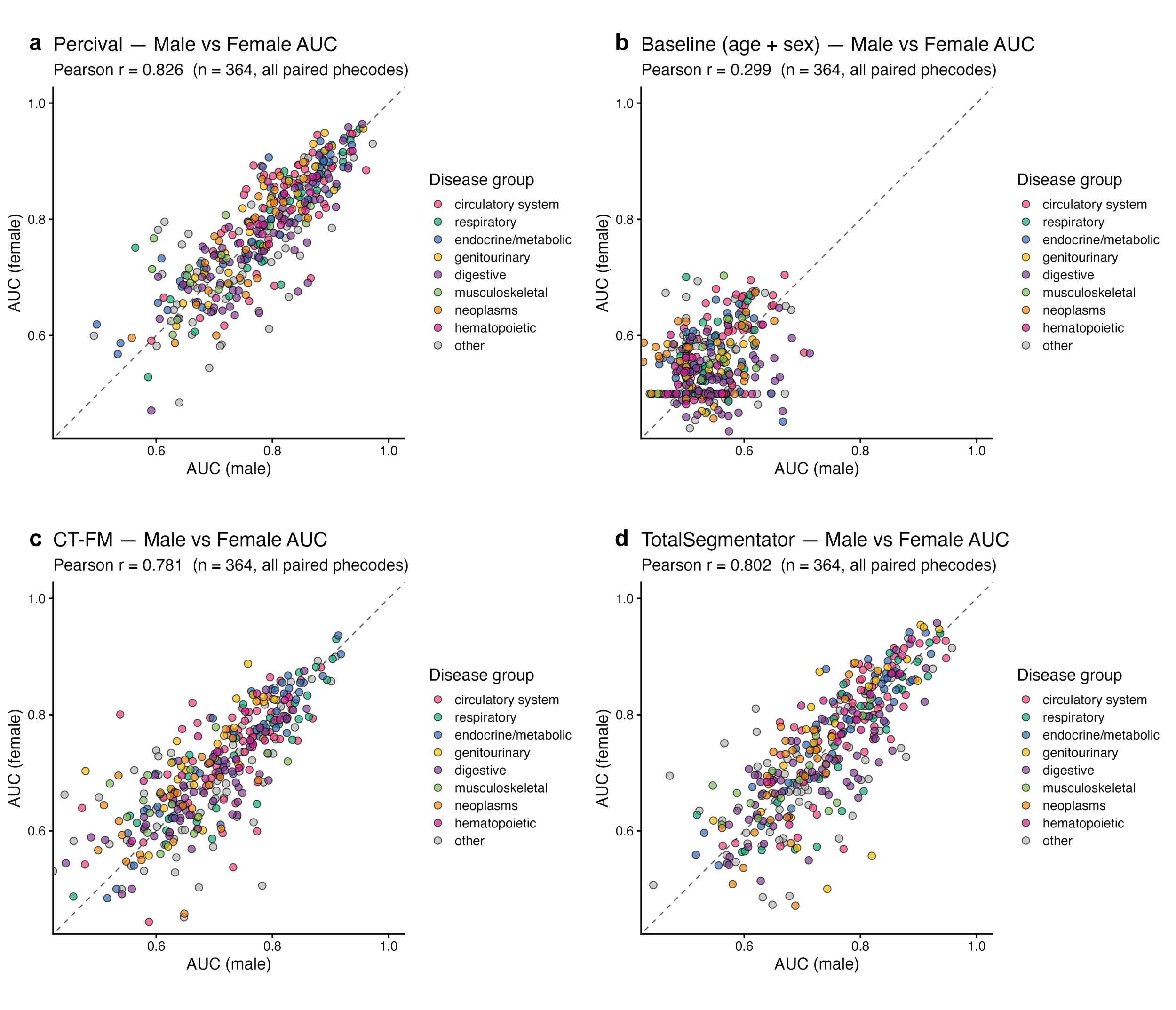

**Figure S12.** Sex-specific replication of disease classification performance in abdomen/pelvis CT, comparing Percival to the demographic baseline, CT-FM, and TotalSegmentator. Each point represents one phecode plotted by area under the receiver operating characteristic curve (AUC) computed separately in male and female participants (n=373 phecodes). Points are colored by disease group, with phecodes outside the eight major groups shown as "other." The dashed diagonal indicates perfect cross-sex concordance. Pearson r is annotated in each panel. (**a**) Percival shows the highest cross-sex concordance (Pearson r=0.826). (**b**) Demographic baseline of age and sex shows the lowest concordance (Pearson r=0.299). (**c**) CT-FM (Pearson r=0.781). (**d**) TotalSegmentator (Pearson r=0.802).

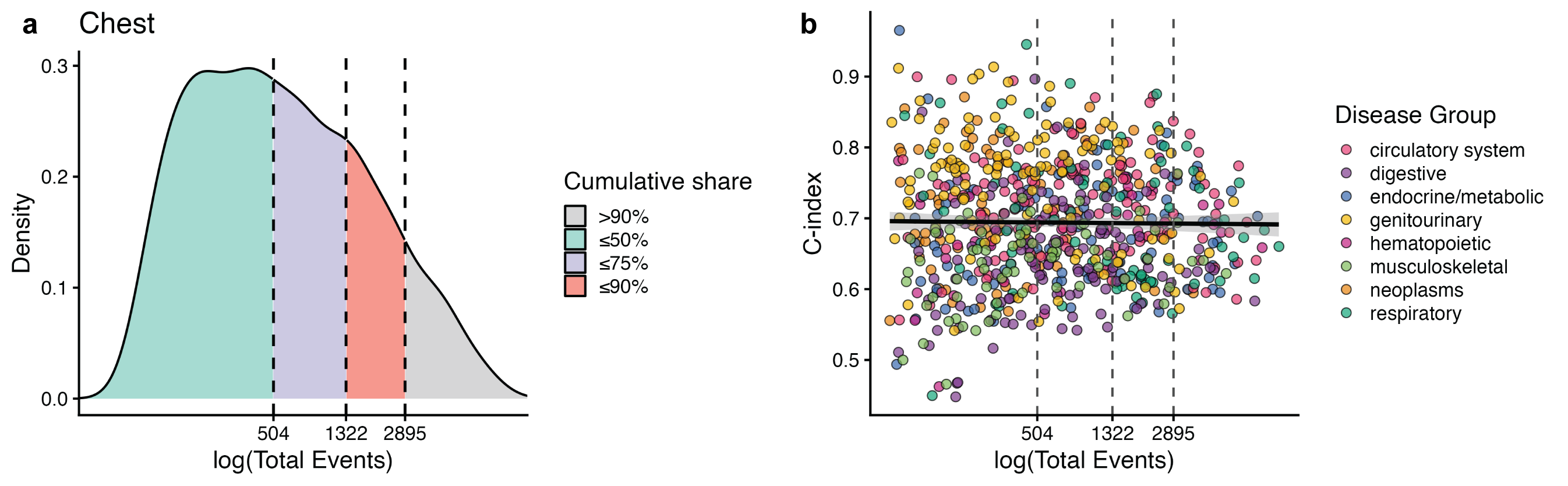

**Figure S13.** Distribution of event counts and Percival's prognostic performance for chest CT-derived predictions. (**a**) Density distribution for 1,150 conditions shaded by cumulative share quartiles. Dashed vertical lines mark the 50th (n=435), 75th (n=1,207), and 90th (n=2,736) percentiles of the event-count distribution. (**b**) Scatter plot of Percival's concordance index (C-index) from chest CT embeddings against the log-transformed number of events for a select group of conditions, with points colored by disease group. The fitted line and shaded band show a linear regression with 95% confidence interval. Vertical dashed lines mark the same percentile thresholds as in panel (**a**).

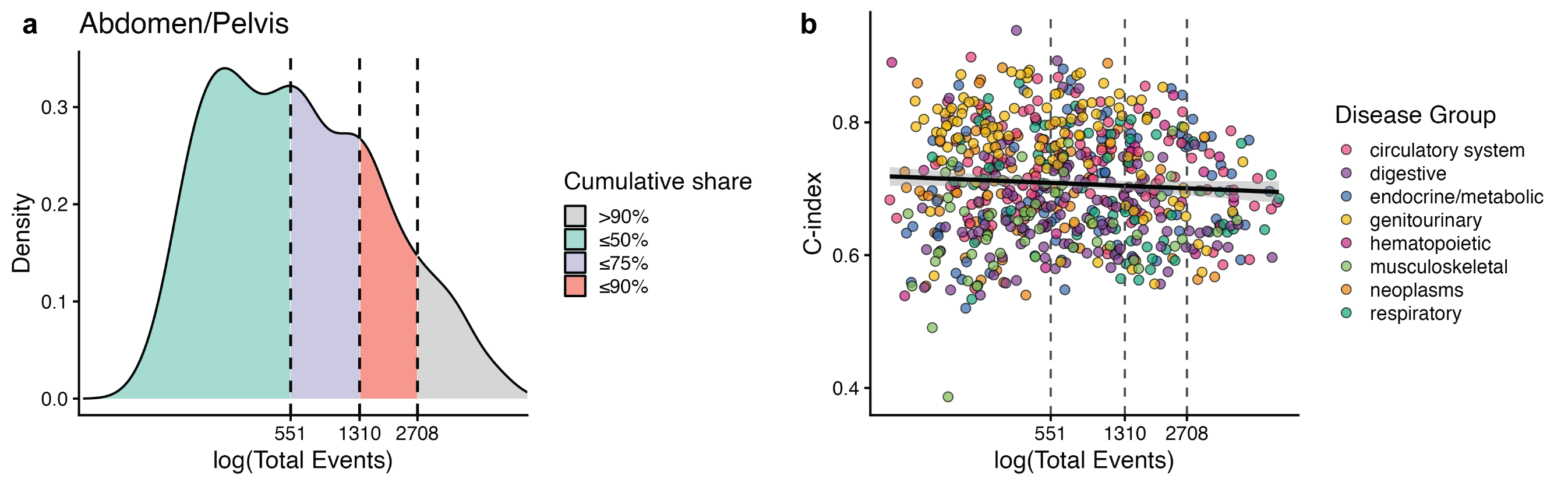

**Figure S14.** Distribution of event counts and Percival's prognostic performance for abdomen/pelvis CT-derived predictions. (**a**) Density distribution for 1,160 conditions shaded by cumulative share quartiles. Dashed vertical lines mark the 50th (n=395), 75th (n=1,077), and 90th (n=2,374) percentiles of the event-count distribution. (**b**) Scatter plot of Percival's concordance index (C-index) from abdomen/pelvis CT embeddings against the log-transformed number of events for a select group of conditions, with points colored by disease group. The fitted line and shaded band show a linear regression with 95% confidence interval. Vertical dashed lines mark the same percentile thresholds as in panel (**a**).

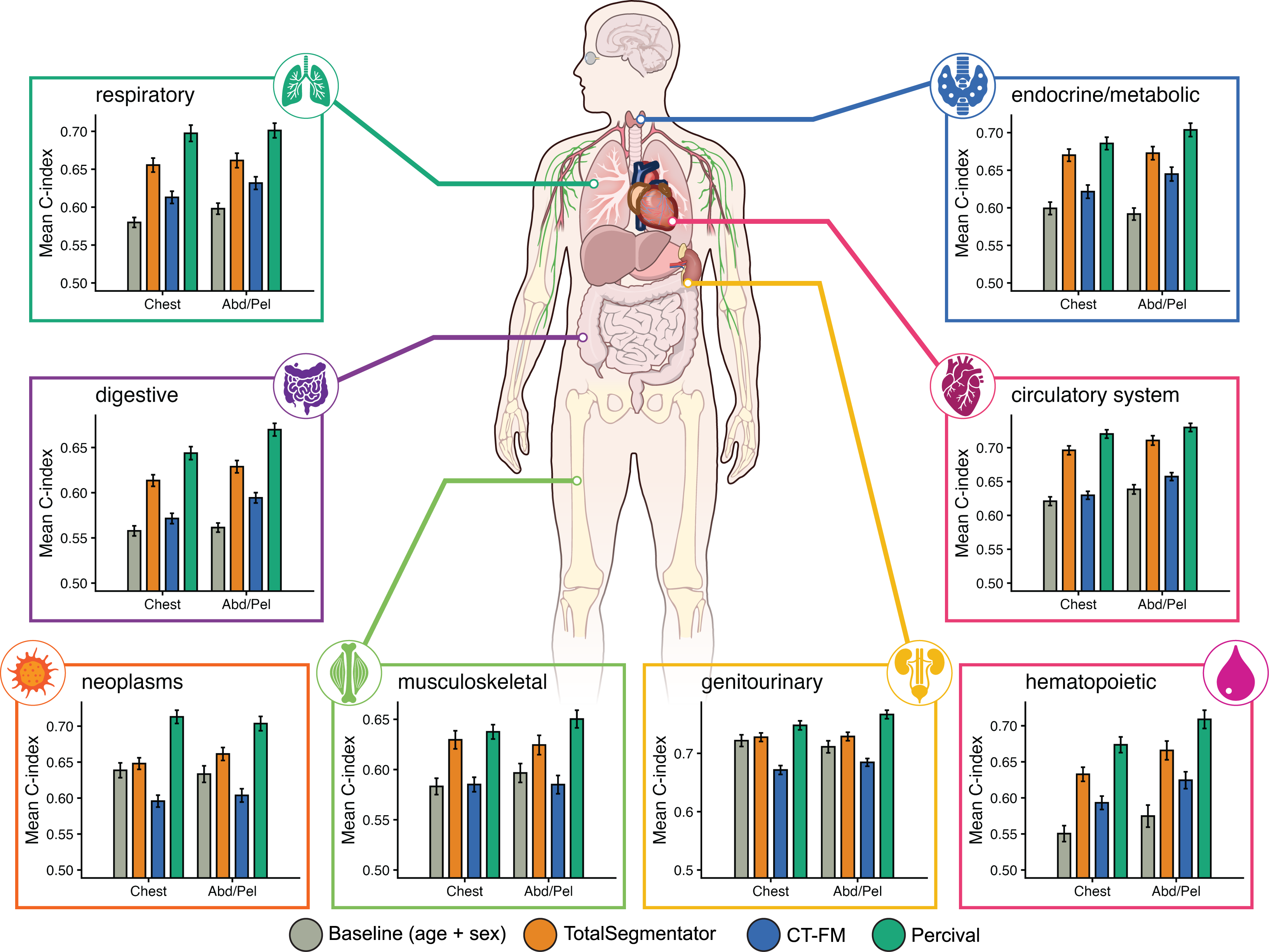

**Figure S15.** Mean concordance index (C-index) across the eight disease groups (circulatory system, respiratory, digestive, genitourinary, musculoskeletal, neoplasms, endocrine/metabolic, hematopoietic) for chest and abdomen/pelvis CT, comparing Percival to the demographic baseline (age + sex), TotalSegmentator, and CT-FM. Error bars represent the standard error of the mean across conditions within each disease group. Human anatomical art provided by NIH BIOART^2^.
